# Genetic susceptibility to Lewy body disease and the distribution of Lewy pathology

**DOI:** 10.64898/2026.10.21.26366146

**Authors:** Lesley Y. Wu, Jared W. Taylor, Miko Valori, Tatiana Georgiades, Tessa du Toit, Ana-Luisa Gil-Martinez, Eleanor J. Stafford, Kristin Levine, Zih-Hua Fang, Cornelis Blauwendraat, Andrew B. Singleton, Sonja W. Scholz, Bryan J. Traynor, John Hardy, Patrick Chinnery, Richard Cain, Claire Troakes, Djordje Gveric, Catriona McLean, Seth Love, Andrew King, Andrew C. Robinson, Federico Roncaroli, Claire Shepherd, Glenda Halliday, Laura Parkkinen, Christopher M Morris, Johannes Attems, Colin Smith, Geidy E. Serrano, Thomas G. Beach, Steve Gentleman, Thomas T. Warner, Tammaryn Lashley, Zane Jaunmuktane, Owen A. Ross, Dennis W. Dickson, Lasse Pihlstrøm, Raquel Real, Huw R Morris, the Global Parkinson’s Genetics Program (GP2)

## Abstract

Genome-wide association studies of Parkinson’s disease and dementia with Lewy bodies largely rely on clinical diagnoses, which can misclassify cases and cannot characterise the anatomical distribution of Lewy pathology. Autopsy cohorts allow identification of variants associated both with developing Lewy body disease and with the neocortical extent of its pathology.

We meta-analysed genome-wide data from 2,661 individuals of European ancestry with autopsy-confirmed Lewy body disease and 12,068 controls (2,124 autopsy-confirmed) from the Global Parkinson’s Genetics Program (GP2) and the National Institutes of Health NeuroBioBank, and estimated genetic correlation with clinically diagnosed Alzheimer’s disease. In 2,309 cases from the GP2 and the Mayo Clinic Jacksonville Brain Bank, we compared neocortical (n = 1,362) with brainstem or limbic (n = 947) Lewy pathology, adjusting for Alzheimer’s disease co-pathology, age at death and sex.

Under a neuropathological case definition, five established loci reached genome-wide significance: *APOE* ε4 (OR 2.27, 95% CI 2.06-2.50), *GBA1* E326K (OR 2.57, 95% CI 2.01-3.28), *SNCA* (OR 1.35, 95% CI 1.23-1.48; *TMEM175* p.M393T (OR 1.30, 95% CI 1.19-1.43), and *HLA* class II (OR 0.69, 95% CI 0.61-0.79). Lead associations differed by antemortem diagnosis. Across all cases, the *SNCA* association was indexed by a 5′ signal previously associated with dementia with Lewy bodies and rapid eye movement sleep behaviour disorder, rather than the canonical 3′ Parkinson’s disease variant. Among 1,216 cases diagnosed during life with Parkinson’s disease, with or without dementia, the 3′ variant was the lead *SNCA* signal, and the *APOE* ε4 effect was attenuated (OR 1.57, 95% CI 1.37-1.81). Genetic correlation with Alzheimer’s disease (0.33, *P* = 0.02) persisted after excluding the *APOE* region (0.31, *P* = 0.03). Within cases, *APOE* ε4 was the only susceptibility locus associated with neocortical involvement (OR 1.70, 95% CI 1.45-1.98; P = 2.5 × 10^−1^^1^), including among 666 GP2 cases without significant Alzheimer’s disease co-pathology (OR 1.76, 95% CI 1.18-2.63; P = 0.005). Each additional ε4 allele was also associated with a more advanced Lewy pathology stage (OR 1.76, 95% CI 1.50-2.06).

Autopsy-confirmed Lewy body disease shares its major susceptibility loci with clinically defined disease. Of these, only APOE ε4 was also associated with neocortical involvement, independently of measured Alzheimer’s disease co-pathology, suggesting that it influences not only whether Lewy pathology develops but how far it extends.

## Introduction

Lewy body disease (LBD) is an umbrella term encompassing Parkinson’s disease (PD), of which PD dementia (PDD) can be considered a subtype, and dementia with Lewy bodies (DLB). These are clinically distinct manifestations of a shared neuropathological process characterised by the accumulation of alpha-synuclein aggregates as Lewy bodies (LBs) and Lewy neurites (LNs). Genome-wide association studies (GWAS) of these clinically defined syndromes have significantly advanced our understanding of the genetic architecture of synucleinopathies ^1,2^. However, the resolution of these studies may be constrained by the accuracy of clinical diagnoses. Clinicopathological studies have demonstrated that 10-15% of individuals diagnosed with PD, PDD or DLB during life are misclassified, with alternative neuropathological diagnoses at autopsy ^3–8^. Therefore, it is possible that some GWAS signals for PD relate to patients with non-synuclein pathologies, and pathologically defined case-control studies are well placed to address this.

Post-mortem assessment overcomes this limitation but reveals an additional layer of heterogeneity. The anatomical distribution of LB pathology varies markedly between individuals, with some showing a predominantly brainstem-focused pattern, while others show extensive involvement of the limbic system and neocortex. Despite advances in identifying genetic risk factors for PD, PDD and DLB, the specific genetic determinants contributing to the distribution of LB pathology remain poorly understood.

The frequent co-occurrence of Alzheimer’s disease (AD) pathology compounds the complexity of the diseases, particularly in patients with extensive neocortical LB pathology. Most individuals with DLB have Aβ pathology, and 30-50% have severe AD neuropathological change ^9,10^. *APOE*ε*4* is a risk allele for AD, and has also been identified as a strong genetic risk factor for developing PDD/DLB ^1^, but has little effect on PD risk in clinically defined case-control studies. Because ε4 promotes Aβ accumulation ^11^, its association with DLB is expected given the prevalence of AD co-pathology; the more informative question is whether it also exacerbates α-synuclein pathology through mechanisms independent of AD pathology.

To address these questions, we performed a genome-wide association meta-analysis of 2,661 neuropathologically confirmed LBD cases and 12,068 controls, in which case status was defined by LB pathology rather than clinical diagnosis. We then compared these associations with GWAS from clinically diagnosed PD. Lastly, we performed a GWAS among LBD cases subset by neuroanatomical disease stage and identified genetic risk factors of neocortical α-synuclein pathology, as opposed to predominantly subcortical pathology.

## Methods

### Participants

Participants were identified from the Global Parkinson’s Genetic Program (GP2), the National Institutes of Health NeuroBiobank (NIHNBB), and the Mayo Clinic Jacksonville Brain Bank. All samples were collected with appropriate informed consent from the participant or their legal representative under protocols approved by their local institution. The present study was approved by the University College London Research Ethics Committee (23473/001).

#### Global Parkinson’s Genetics Program

GP2 is an initiative which aims to genotype over 250,000 participants and whole genome sequence (WGS) over 100,000 individuals to further our understanding of the genetic architecture of PD ^12^. Individuals with a post-mortem diagnosis of LBD made by the contributing brain banks, neurologically normal controls and clinical controls were selected from GP2 release 11. All were of genetically inferred European ancestry ^13^, and had recorded clinical sex and either age at death or age at study participation (for the clinical controls). A subset of cases also had Unified Staging System for Lewy body Disorders (USSLB) ^14^, Braak LB ^15^ stage or McKeith stage ^16^, Braak neurofibrillary tangle (NFT) stage ^17^ and Thal Phases ^18^. Samples were genotyped using the Illumina Global Diversity Array chip with Neurobooster (NBA) ^19^, called from IDAT files and quality-controlled with GenoTools, and imputed against the Trans-Omics for Precision Medicine (TOPMed) r2 panel on the TOPMed Imputation Server (https://imputation.biodatacatalyst.nhlbi.nih.gov/; Eagle v2.4 phasing, Minimac4). These imputed NBA data were used for the GWAS. Brain bank samples also underwent WGS. Alignment and small variant calls against the GRCh38DH reference genome were produced. The Broad Institute’s joint discovery pipeline was used to perform joint genotyping across all samples that meet WGS quality control criteria, as outlined by the AMP-PD (https://amp-pd.org). High-quality variants were retained post-joint-genotyping if they met the following thresholds: a PASS flag after variant quality score recalibration, a call rate >0.95, genotype quality >20, read depth >5, and heterozygous allele balance between 0.25 and 0.75. We used the WGS data for the HLA allele analysis.

#### National Institute of Health Neurobiobank

NIHNBB samples were contributed by six brain tissue repositories. The NIHNBB database (https://neurobiobank.nih.gov/) was queried to identify individuals with an autopsy-confirmed diagnosis of LBD or neurologically normal controls. Individuals with available WGS data and genetically inferred European ancestry (as determined using Peddy^20^) were subsequently selected for inclusion. WGS was performed on the Illumina NovaSeq 6000 system. Further details on the cohort and sequencing methods are provided in Hupalo et al., 2026 ^21^.

#### Mayo Clinic Jacksonville Brain Bank

A detailed description of case recruitment and selection from the Mayo Clinic Jacksonville Brain Bank for Neurodegenerative Disorders can be found in Tunold et al. ^22^. Briefly, all included cases had a post-mortem diagnosis of LBD, were Caucasian, and were unrelated. Braak LB stages, Braak NFT stages and Thal phases were available for all patients. Samples were genotyped on the Illumina Infinium OmniExpress-24 array. Standard quality control was performed and variant imputation against the Haplotype Reference Consortium was carried out.

### Quality Control of Genetic data

We performed additional QC steps on the imputed and sequenced data. Single nucleotide polymorphisms (SNP) were excluded if they deviated significantly from Hardy-Weinberg equilibrium (P < 1 ×10^−8^), had a minor allele frequency less than 1%, or were located on non-autosomal chromosomes (X, Y, or mitochondrial). We also excluded related individuals (kinship coefficient > 0.088, i.e. second-degree relatives or closer) from analysis.

### Harmonization of pathological data

As different neuropathological staging systems have been developed over time, the participating brain banks used different classification schemes and did not uniformly collect all pathological measures. Therefore, a harmonization strategy was applied to maximise the number of samples available for analysis while maintaining consistency across cohorts. For LB pathology, a synthetic neocortical - subcortical endophenotype was generated by harmonizing categorical and ordinal staging systems. Cases with neocortical involvement, defined as USSLB/McKeith neocortical stage or Braak LB ≥ 5, were classified as neocortical. Cases without neocortical LBs, defined as USSLB/McKeith brainstem or limbic stage or Braak LB stages 1 - 4, were classified as subcortical (Supplementary Table 1). The presence of AD co-pathology was assessed using an adaptation of the NIA-AA ABC Score, as described by Tunold *et al.* (2023) ^22^. In brief, Thal amyloid phases and Braak NFT stages were combined to generate a dichotomous measure of AD co-pathology. Cases were classified as having no significant AD co-pathology if they were assigned a “no” or “low” score on both staging systems, whereas cases were classified as having significant AD co-pathology if they received an “intermediate” or “high” score (Supplementary Table 2).

### Statistical analysis

#### Single-variant association analyses

We performed two main GWAS, the first to identify common genetic variants associated with susceptibility to autopsy-confirmed LBD, and the second to identify genetic factors associated with the anatomical distribution of LB pathology. Association testing was performed using PLINK 2.0 ^23^. The number of genetic principal components (PCs) included as covariates was determined by inspection of scree plots for each analysis.

The first GWAS compared autopsy-confirmed LBD cases with controls. Logistic regression was performed independently in the GP2 and NIHNBB cohorts, adjusting for age, sex, and PC1-PC5, where age was defined as age at death for the autopsy-confirmed cases and age at participation for the controls. AD co-pathology was not included as a covariate in the case-control analysis, as the aim was to estimate the total genetic contribution to pathologically defined LBD, including effects mediated through AD pathology; AD-independent effects were assessed separately in the within-case analyses. Cohort-level summary statistics were subsequently combined by meta-analysis. Genomic inflation was assessed using the genomic control factor and quantile-quantile plots (Supplementary Figure. 1). As a sensitivity analysis, we restricted cases to individuals with autopsy-confirmed LBD and an antemortem clinical diagnosis of PD or PDD, i.e. excluding clinically diagnosed DLB. This analysis was confined to the GP2 cohort, the only cohort with available antemortem diagnostic information, and comprised 1,216 cases (890 PD, 326 PDD) and 10,517 controls (Supplementary Table 3). The same model specification was used.

The second GWAS tested associations with the anatomical distribution of LB pathology among LBD cases, dichotomised as neocortical or subcortical. Cases came from GP2 and the Mayo Clinic Jacksonville Brain Bank; NIHNBB was excluded because harmonised LB and AD staging was available for only 139 individuals. Of the 1,928 GP2 cases in the primary analysis, 1,331 (69.0%) had sufficient staging; with 978 Mayo cases, this gave 2,309 cases (1,362 neocortical, 947 subcortical; Table 5). Logistic regression was performed in each cohort, adjusting for age at death, sex, AD co-pathology and PC1-PC3. Pathological stage was also analysed as an ordinal trait (brainstem-predominant, limbic, diffuse neocortical) by proportional-odds regression on OrdinalGWAS ^24^ for Julia, with the same covariates. The proportional-odds assumption was assessed in each cohort with the Brant test at the lead variant of each locus with *P* < 5 × 10^−6^, and per-cohort results were combined by fixed-effect inverse-variance meta-analysis. Two sensitivity analyses examined the case-case lead variant. In 1,034 cases with recorded disease duration, it was re-estimated with sex and principal components alone, then with age at death, AD co-pathology and disease duration added singly and jointly; comparing this base model with the primary estimate separated the effect of the covariates from that of the restricted sample. It was also tested within strata of AD co-pathology, and the stratum estimates compared by a z-test of the difference in log odds ratios. To test whether the case-control loci also influenced distribution, their lead variants were evaluated in the case-case results against a Bonferroni-corrected threshold. For loci without an association, power at that two-sided threshold was calculated for a true odds ratio equal to the case-case lead-variant estimate, using each variant’s own standard error.

For both main GWAS, meta-analysis was performed using METAL ^25^ with a fixed-effect inverse-variance-weighted model. Variants were retained if present in all contributing cohorts and if the range of allele frequencies across cohorts was <15%. Between-cohort heterogeneity was assessed using Cochran’s Q test and the I² statistic, which estimates the proportion of variation in effect estimates across cohorts attributable to heterogeneity rather than chance. Variants demonstrating substantial heterogeneity (Q P < 0.05 and I² >80%) were excluded. Genome-wide significance was defined as P < 5 × 10^−8^ and suggestive significance as P < 5 × 10^−6^.

Secondary independent signals were identified by conditional and joint analysis in GCTA-COJO (v1.93.0 beta, default settings)^26^, with linkage disequilibrium (LD) estimated from unrelated GP2 controls of genetically inferred European ancestry.

#### Functional annotation and gene-based association

Summary statistics from the autopsy-confirmed LBD meta-analysis (N = 14,729) were processed with FUMA v1.8.2 (SNP2GENE), using the 1000 Genomes Phase 3 European reference panel and Ensembl v102 protein-coding gene definitions. Independent significant SNPs were defined as P < 5 × 10^−8^ and r² < 0.6, lead SNPs as r² < 0.1, and candidate SNPs as those in linkage disequilibrium (r² ≥ 0.6) with an independent significant SNP and P < 0.05. The extended MHC was excluded from locus definition and gene mapping and retained for annotation only. Gene-based association was performed with MAGMA v1.08 as implemented in FUMA, using the SNP-wise mean model with no flanking window.

#### Classical HLA allele analysis

Classical *HLA* alleles were called from whole-genome sequence alignments using DRAGEN, which reports calls at four-field resolution; these were truncated to two fields for analysis. Analysis was restricted to individuals with sequence data passing quality control and non-missing case-control status and genotype-inferred sex. One member of each related pair (kinship ≥ 0.088) was excluded, giving 1,051 cases and 2,626 controls (1,986 male, 1,691 female).

Of 1,294 alleles called across 39 loci, calls supported by a mean read depth below 90 were discarded, leaving 1,272 alleles, of which 165 were carried by more than 5% of individuals and observed in both cases and controls. The 72 alleles at the seven loci (*HLA-A*, *-B*, *-C*, *-DPB1*, *-DQA1*, *-DQB1* and *-DRB1*) were pre-specified as the inference set; the remaining 93, at other MHC-region loci, were treated as exploratory.

Each allele was tested against case-control status by two-sided logistic regression, entering allele dosage with sex and the first five genetic principal components, computed across all individuals. Testing was performed on complete cases at each locus; individuals with a missing call at a given locus were excluded from tests of alleles at that locus. Alleles were coded additively (0/1/2), dominantly (0/1/1) and recessively (0/0/1). The false discovery rate was controlled by the Benjamini–Hochberg procedure across the classical-locus alleles within each encoding.

#### Genetic correlation and cross-disease comparison

To place the genetic architecture of neuropathologically confirmed LBD in the context of clinically defined neurodegenerative disease, we estimated genome-wide genetic correlations with GWAS of clinical PD ^2^, clinical DLB ^1^, and AD ^27,28^ using linkage disequilibrium score regression (LDSC) ^29^. LDSC was applied to summary statistics under default settings. AD genetic correlations were estimated using both the IGAP ^27^ and the larger Bellenguez *et al.* dataset incorporating proxy-defined cases ^28^, and repeated after excluding the *APOE* region (chr19:43,900,000 - 46,000,000, GRCh38) to assess the contribution of the *APOE* locus.

Overlap with clinical PD at the level of individual loci was assessed by comparing per-allele effect estimates at the independent lead SNPs reported in the GP2 European-ancestry PD meta-analysis ^2^. Lead SNPs were extracted from the pathological LBD summary statistics, ambiguous palindromic variants were excluded, and effect estimates were oriented to the PD risk-increasing allele. Concordance was summarised by Pearson correlation of the two sets of effect estimates, by the proportion of variants with concordant direction of effect, and by a beta-beta plot.

Differences in effect size between studies were formally tested using a two-sample z-statistic, with two-sided P values derived from the standard normal distribution. This test assumes independence of the two estimates; as GP2 cohorts contributed to both meta-analyses, the resulting P values are conservative with respect to the shared component of the samples. Significance was defined by Bonferroni correction for the 134/157 available variants tested (P < 3.7 × 10^−4^).

## Results

### Autopsy-confirmed LBD case-control meta-analysis identifies five genome-wide significant loci

To define the common-variant architecture of LBD, we performed meta-analysis across two neuropathologically confirmed cohorts. All 2,661 cases carried an autopsy diagnosis of LBD (Global Parkinson’s Genetic Program [GP2] n = 1,928; National Institutes of Health NeuroBiobank [NIHNBB]; n = 733), and were compared to 12,068 controls, including 2,124 autopsy-confirmed neurologically normal controls (GP2 n = 9,944 clinical controls and 573 autopsy-confirmed controls; NIHNBB 1,551 autopsy confirmed controls; Table 1). Pathologically diagnosed LBD encompassed a range of clinical diagnoses, including PD, PDD, DLB, AD and others, reflecting the phenotypic heterogeneity of the disease.

**Table 1.** Demographic and clinicopathological characteristics of autopsy-confirmed LBD cases and of autopsy-confirmed and clinical controls.

| Cohort | Clinical diagnosis | PD | PDD | DLB | AD | Other | Unknown | Controls |
| --- | --- | --- | --- | --- | --- | --- | --- | --- |
|  | Pathological diagnosis | LBD | LBD | LBD | LBD | LBD | LBD | - |
| GP2 | N | 890 | 326 | 194 | 208 | 253 | 57 | 10,517# |
|  | Sex, F (%) | 342 (38.4%) | 94 (28.8%) | 37 (19.1%) | 91 (43.8%) | 97 (38.3%) | 23 (40.4%) | 5,577 (53.0%) |
|  | Age<sup>^</sup>, mean (SD) | 78.8 (7.6) | 77.8 (6.5) | 78.3 (7.7) | 81.2 (9.4) | 79.1 (9.5) | 81.5 (9.3) | 63.4 (14.0) |
|  | Presence of AD pathology* | 240/612 (39.2%) | 113/290 (39.0%) | 92/155 (59.4%) | 171/190 (90.0%) | 113/185 (61.1%) | 2/2 (100.0%) | - |
| NIHNBB | N | 247 | 69 | 160 | 98 | 126 | 33 | 1,551 |
|  | Sex, F (%) | 72 (29.1%) | 21 (30.4%) | 48 (30.0%) | 51 (52.0%) | 57 (45.2%) | 17 (51.5%) | 581 (37.5%) |
|  | Age at death, mean (SD) | 77.9 (7.7) | 79.7 (7.4) | 76.2 (7.8) | 79.3 (9.9) | 78.3 (10.7) | 77.2 (6.6) | 59.0 (19.6) |
|  | Presence of AD pathology* | 14/36 (38.9%) | 19/34 (55.9%) | 27/46 (58.7%) | 39/44 (88.6%) | 15/34 (44.1%) | NA | - |

\* % calculated on available data: N = 1,434 for GP2, 194 for NIHNBB.

The meta-analysis identified five genome-wide significant loci (P < 5 × 10^−8^; Figure 1, Table 2), all corresponding to previously reported clinically defined PD or DLB GWAS associations ^1,2^. The strongest association was at *APOE*, indexed by the ε4-defining variant rs429358 (OR = 2.27, 95% CI 2.06-2.50, P = 1.0 × 10^−61^), followed by *GBA1*, where the lead variant was the missense E326K (rs2230288; OR = 2.57, 95% CI 2.01-3.28, P = 5.4 × 10^−14^). The *SNCA* signal was indexed by the intronic variant rs2737030 (OR = 1.35, 95% CI 1.23-1.48, P = 2.3 × 10^−10^) and *TMEM175* by p.M393T (rs34311866; OR = 1.30, 95% CI 1.19-1.43, P = 5.6 × 10^−9^). The fifth association mapped to the *HLA* class II region (rs9275382; OR = 0.69, 95% CI 0.61-0.79, P = 7.3 × 10^−9^). All five lead variants showed a consistent direction of effect in both cohorts, with no significant between-cohort heterogeneity (all P het > 0.05), although the *HLA* association was driven mainly by the GP2 cohort.

**Figure 1.**
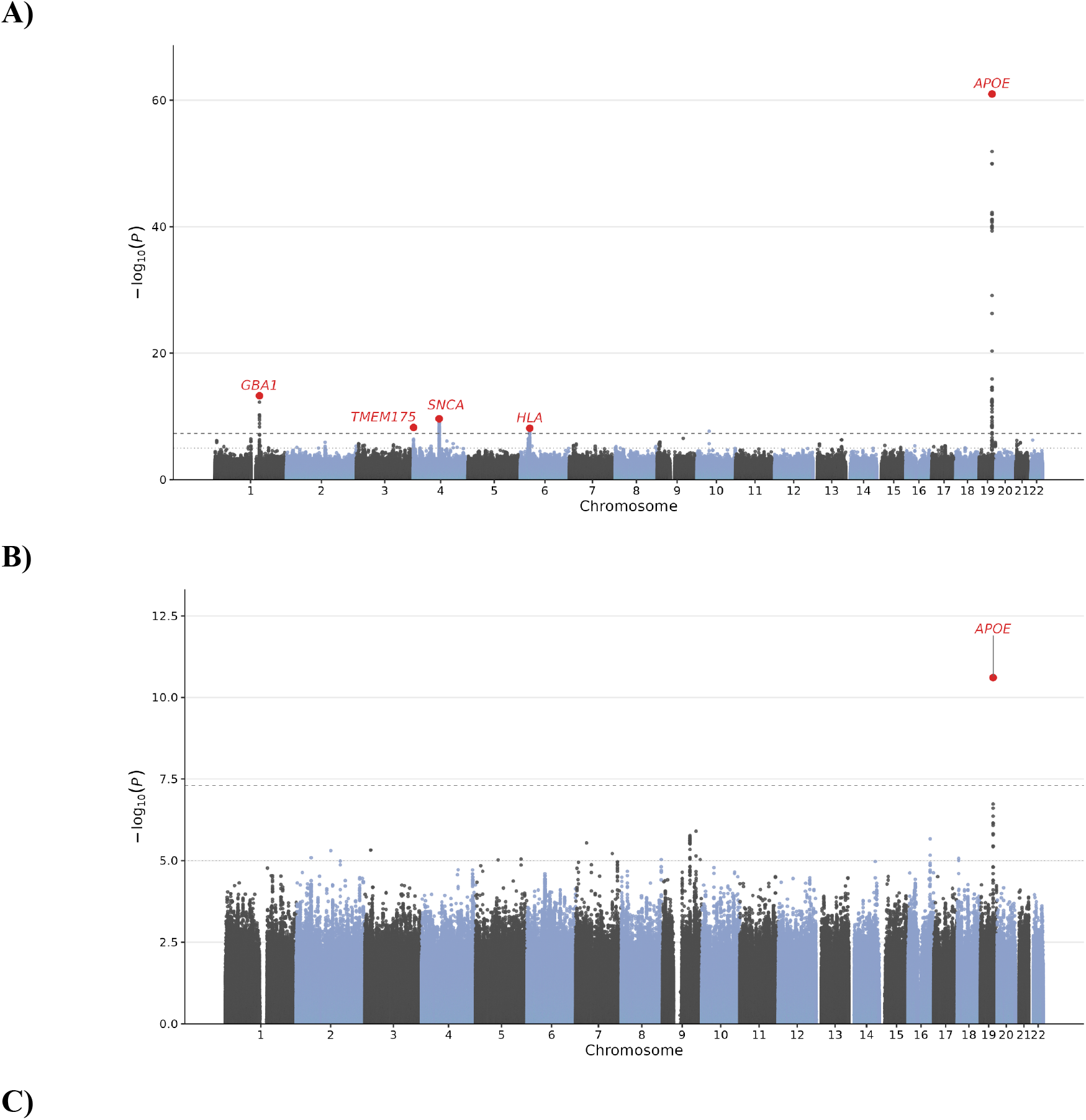
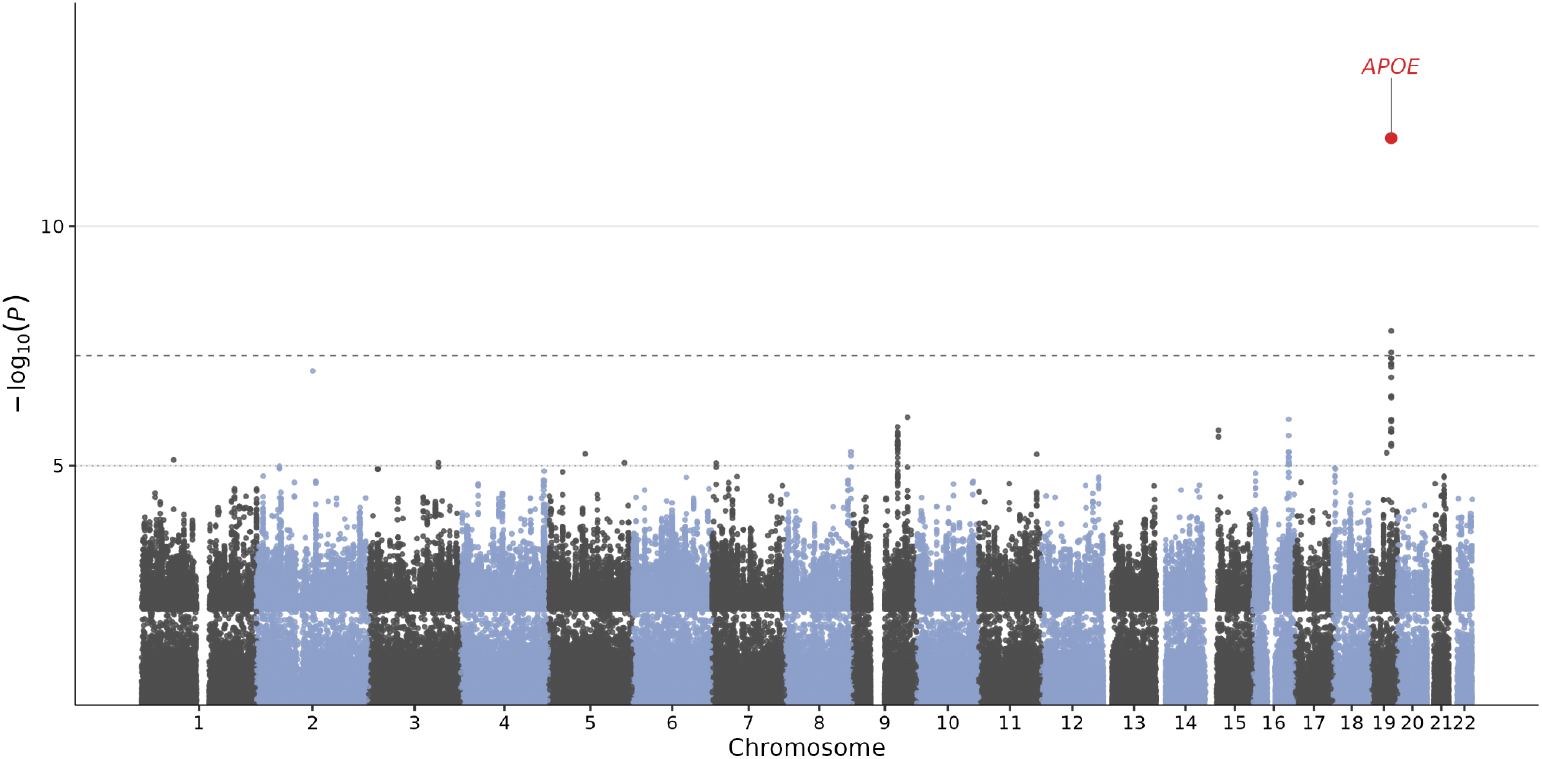
Manhattan plot of the genome-wide association meta-analysis of neuropathologically confirmed Lewy body disease. A) Case-control meta-analysis (*n =* 2,661 Lewy body disease cases and 12,068 controls). Five loci reached genome-wide significance; these are highlighted in red and labelled by candidate gene: *GBA1* (1q22), *TMEM175* (4p16.3), *SNCA* (4q22), *HLA* class II (6p21.32) and *APOE* (19q13.32). B) Case-case meta-analysis comparing cortical with subcortical Lewy body distribution (*n =* 1,362 neocortical and 947 subcortical cases), in which only *APOE* (19q13.32) reached genome-wide significance. C) Ordinal meta-analysis of Lewy body pathological stages (brainstem, transitional/limbic, diffuse neocortical) under a proportional-odds model in the same 2,309 cases as B), in which again, *APOE* (19q13.32) reached genome-wide significance, indexed by rs429358. In all panels, the −log₁₀-transformed *P value* is plotted on the y-axis against chromosomal position on the x-axis, and the dotted line indicates the genome-wide significance threshold (*P =* 5 × 10⁻⁸).

**Table 2.**
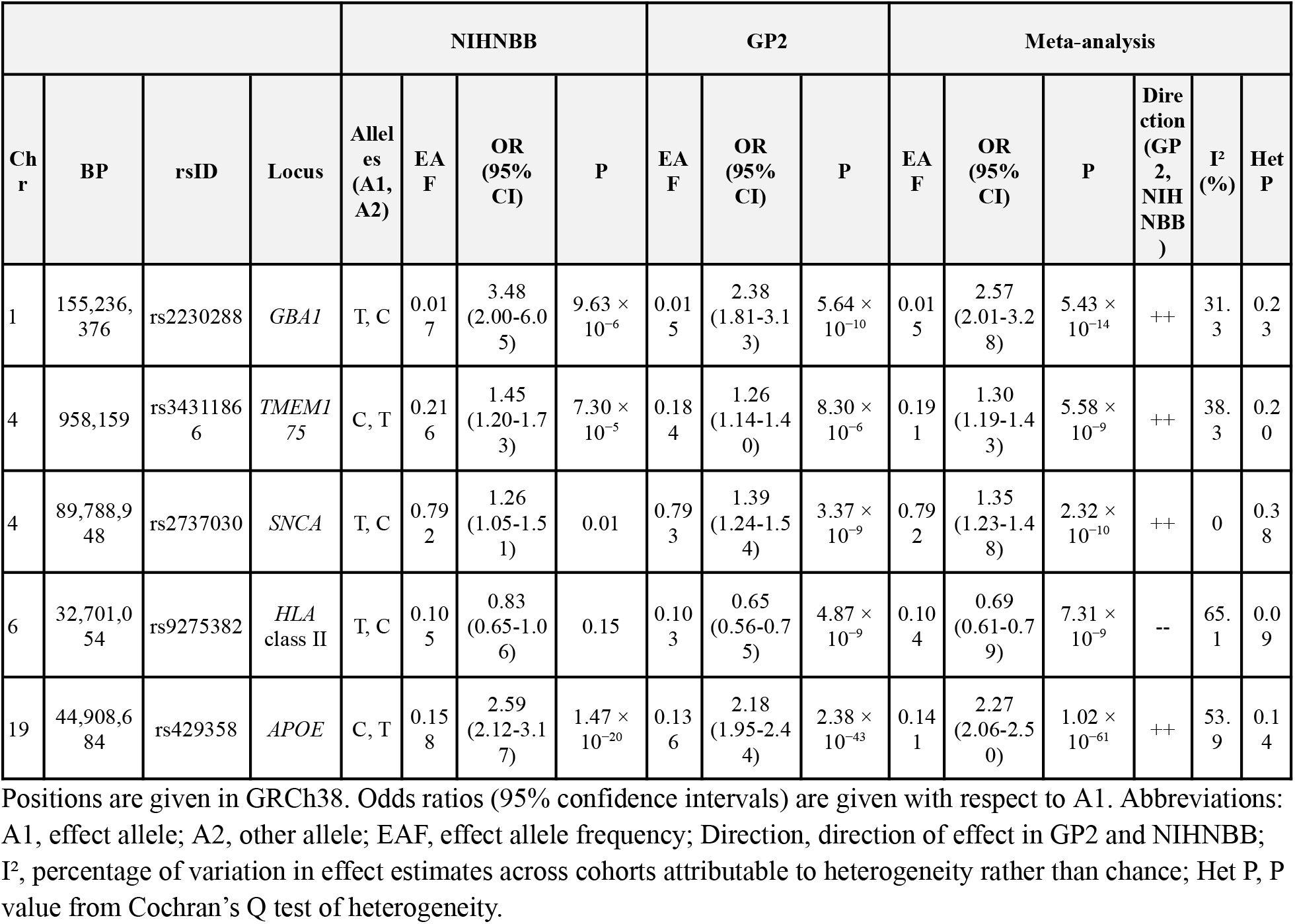
Genome-wide significant lead SNPs from the neuropathologically confirmed Lewy body disease meta-analysis.

To assess whether these associations were influenced by the inclusion of clinically diagnosed DLB cases, we repeated the GWAS after restricting cases to individuals with antemortem diagnosis of PD or PDD and neuropathologically confirmed LBD. This analysis was confined to GP2 and comprised 1,216 cases (890 PD, 326 PDD) and 10,517 controls (Supplementary Table 3). Four loci reached genome-wide significance: *GBA1* (rs2230288, OR = 2.80, 95% CI 2.07-3.77, P = 1.5 × 10^−11^), *SNCA* (rs356182, OR = 1.35, 95% CI 1.22-1.49, P = 2.7 × 10^−9^), the extended *MHC* class I region (rs3117443, OR = 0.74, 95% CI 0.67-0.81, P = 7.8 × 10^−10^) and *APOE* (rs429358, OR = 1.57, 95% CI 1.37-1.81, P = 1.2 × 10^−10^; Supplementary Table 4, Supplementary Figure 2). Compared with all GP2 cases with neuropathologically confirmed LBD, irrespective of antemortem diagnosis, the *APOE* effect was smaller (OR = 1.57 versus 2.18). At *SNCA* and the *MHC*, the lead variant changed. At both loci the two variants are independent signals, and the change reflected a shift in their relative strength rather than re-indexing of a single signal (PD/PDD cases versus all GP2 cases). At *SNCA*, the effect of rs356182 was larger (OR = 1.35 versus 1.17) and that of rs2737030 smaller (1.29 versus 1.39). At the *MHC*, the effect of rs3117443 was further from the null (0.74 versus 0.81), whereas that of rs9275382 was essentially unchanged (0.68 versus 0.65), so its loss of genome-wide significance reflects the smaller sample rather than attenuation. The two MHC variants were in linkage equilibrium (r² < 0.001, D′ = 0.09), and each remained associated after conditioning on the other (rs3117443, P = 3.0 × 10^−9^; rs9275382, P = 1.1 × 10^−5^). Effects at the lysosomal loci were similar (*GBA1* 2.80 versus 2.38; *TMEM175* 1.25 versus 1.26); *TMEM175* did not reach genome-wide significance, again consistent with reduced power.

Given the extensive linkage disequilibrium across the *APOE* locus, we next performed stepwise conditional and joint analysis using GCTA-COJO^26^ with LD estimated from GP2 European controls. A single independent signal was retained, and no variant remained genome-wide significant after conditioning on rs429358 (smallest P = 4.9 × 10^−4^).

### At SNCA, the association is indexed by the 5′ signal shared with DLB and REM sleep behavior disorder rather than the canonical 3′ PD signal

To determine whether the association at *SNCA* corresponded to the known PD case-control risk PD locus, we compared the lead variant rs2737030 from our meta-analysis (all clinical diagnoses) with rs356182, a well-established index variant for the canonical PD association at this locus, and the lead variant in the clinically diagnosed, pathologically confirmed PD group ^30^. The variant rs2737030 lies within intron 4 of *SNCA*, approximately 84 kb 5′ of rs356182 (Supplementary Figure 3). The two variants were in low linkage disequilibrium in the GP2 European reference panel (r² = 0.036, D′ = 0.51). rs356182 was only nominally associated in the present meta-analysis (OR = 1.12, 95% CI 1.04-1.21, P = 3.2 × 10^−3^), and conditioning on it left rs2737030 genome-wide significant and essentially unchanged (conditional P = 6.7 × 10^−9^), as the low correlation between them would predict.

We next examined the lead *SNCA* variants previously reported for clinical DLB (rs7680557) ^1^ and REM sleep behaviour disorder (RBD; rs3756059)^31^ GWAS, which lie approximately 50 kb further 5′ of rs2737030 (53 and 47 kb, respectively) and are in near-complete linkage disequilibrium with one another (r² = 0.974, D′ = 0.992). Both variants were genome-wide significant in our meta-analysis (rs7680557, OR = 1.24, P = 8.8 × 10^−9^; rs3756059, OR = 1.23, P = 1.4 × 10^−8^), with concordant effects across contributing cohorts. Both were moderately correlated with rs2737030 (r² = 0.245 and 0.240, respectively; D′ = 0.950 and 0.945), but only weakly correlated with rs356182 (r² = 0.165 and 0.169, D′ = 0.55 and 0.57, respectively).

In contrast to rs356182, conditioning on either 5′ variant attenuated rs2737030 below genome-wide significance (P = 2.3 × 10^−4^ in both cases), and the reciprocal analyses attenuated rs7680557 and rs3756059 to a comparable degree (P = 4.5 × 10^−3^ and P = 6.3 × 10^−3^). Stepwise selection retained a single independent signal at this locus, with no further variant reaching genome-wide significance after conditioning (Supplementary Fig. 2). Together these findings indicate that the *SNCA* association with neuropathologically confirmed LBD is not indexed by the canonical clinically diagnosed PD case-control variant rs356182, which was only nominally associated here, but is instead is identical to the 5′ associations reported in clinical LBD and RBD, most likely representing the same underlying signal.

### Classical HLA alleles at the class II locus

To refine the class II association identified in the primary GWAS, we called two-field classical HLA alleles from whole-genome sequencing data available for a subset of the GP2 cohort (1,051 cases and 2,626 controls). Two of the 72 alleles tested at loci HLA-A, -B, -C, -DPB1, -DQA1, -DQB1 and -DRB1, survived correction, both in the protective direction and both under an additive model: *HLA-DQB1*\*03:02 (OR = 0.72, 95% CI 0.59-0.87, P = 5.4 × 10^−4^, FDR = 0.02) and *HLA-DQA1*\*03:01 (OR = 0.71, 95% CI 0.58-0.86, P = 5.6 × 10^−4^, FDR = 0.02), concordant in direction with the lead variant rs9275382 (Supplementary Table 5). These two alleles encode the α and β chains of the DQ8 heterodimer and are in strong linkage disequilibrium in our cohort (r² = 0.84; 89.1% of *DQB1*\*03:02 carriers also carry *DQA1*\*03:01), and therefore likely represent a single signal rather than two independent associations.

### Gene-based association

To complement single-variant analysis, we performed gene-based association testing and functional annotation of the associated loci on FUMA ^32^. Gene-based analysis identified significant associations at genes within established AD and PD GWAS loci, including *APOC1*, *SNCA*, *TOMM40*, and *APOE* (Bonferroni-corrected P < 2.6 × 10^−6^; Supplementary Table 6). Beyond the established loci, no genes reached the gene-based significance, although a suggestive signal was observed at the 17q21.31 *MAPT* locus (P = 8.5 × 10^−5^).

### Shared genetic architecture with clinical dementia with Lewy bodies, Parkinson’s disease and Alzheimer’s disease

To place the genetic architecture of neuropathologically confirmed LBD in the context of clinically defined neurodegenerative diseases, we estimated genome-wide genetic correlations with GWAS in clinical DLB ^1^, PD ^2^ and AD ^27,28^ using linkage disequilibrium score regression (LDSC) ^29^.

Pathologically confirmed LBD (our results) showed a strong genetic correlation with clinically diagnosed DLB (rg = 1.19, SE = 0.42, P = 4.6 × 10^−3^), as well as clinically diagnosed PD (rg = 1.11, SE = 0.20, P = 3.9 × 10^−8^), consistent with extensive sharing of common genetic risk between pathological LBD and clinical PD, PDD and DLB (Table 3). Cross-trait LDSC intercepts indicated sample overlap with both the clinical PD GWAS^2^ (0.069, SE 0.005) and the DLB GWAS of Chia et al.^1^(0.042, SE 0.005; Table 3). Genetic correlation estimates are robust to such overlap, but these comparisons indicate concordance rather than independent replication.

**Table 3.** LDSC genetic correlations between autopsy-confirmed LBD and related traits.

| Trait | Source | $r_g$ | SE | Z | P | $h^2$<br>(observed) | SE | $h^2$<br>intercept | SE | gcov<br>intercept | SE |
| --- | --- | --- | --- | --- | --- | --- | --- | --- | --- | --- | --- |
| Clinical Parkinson's disease | Leonard et al. 2025 | 1.11 | 0.20 | 5.49 | $3.91 \times 10^{-8}$ | 0.0347 | 0.0027 | 1.0864 | 0.0115 | 0.0686 | 0.0053 |
| Lewy body dementia | Chia et al. 2021 | 1.19 | 0.42 | 2.84 | $4.57 \times 10^{-3}$ | 0.1769 | 0.0809 | 1.0025 | 0.0070 | 0.0421 | 0.0050 |
| Alzheimer's disease | Kunkle et al. 2019 | 0.33 | 0.14 | 2.40 | 0.02 | 0.0732 | 0.0112 | 1.0284 | 0.0089 | 0.0120 | 0.0049 |
| Alzheimer's disease with proxy cases | Bellenguez et al. 2022 | 0.43 | 0.13 | 3.33 | $8.57 \times 10^{-4}$ | 0.0216 | 0.0021 | 1.0679 | 0.0104 | 0.0118 | 0.0052 |
| Alzheimer's disease without *APOE* | Kunkle et al. 2019 | 0.31 | 0.14 | 2.22 | 0.03 | 0.0662 | 0.0093 | 1.0243 | 0.0071 | 0.0111 | 0.0045 |
| Alzheimer's disease with proxy cases without *APOE* | Bellenguez et al. 2022 | 0.42 | 0.13 | 3.28 | $1.00 \times 10^{-3}$ | 0.0213 | 0.0021 | 1.0628 | 0.0089 | 0.0106 | 0.0050 |

Genetic correlations were estimated by linkage disequilibrium score regression (LDSC). Analyses “without *APOE*” exclude the *APOE* region (chr19:43,900,000–46,000,000, GRCh38). Abbreviations: $r_g$ , genetic correlation; SE, standard error; $h^2$ , SNP heritability; gcov intercept, cross-trait LDSC intercept, which reflects sample overlap.

To characterise this overlap at the level of individual risk loci, we compared per-allele effect sizes of the 157 independent signals identified in the 2025 GP2 clinical PD GWAS to their effects in our LBD GWAS (Figure 2). Of these, 134 signals were present and allele-alignable in our LBD dataset and were taken forward for comparison, with all variants oriented to the PD risk-increasing allele. Effect estimates were correlated in the two studies (Pearson’s r = 0.78, P = 1.8 × 10^−28^), and 103 out of 134 signals (77%) showed concordant directions of effect. Most signals lay close to the line of equality, with more falling below it than above, mainly among weaker PD signals, indicating broadly comparable per-allele effects in the two studies, and eight variants at seven loci reached Bonferroni-corrected significance in the present LBD GWAS (P < 3.7 × 10^−4^).

**Figure 2.**
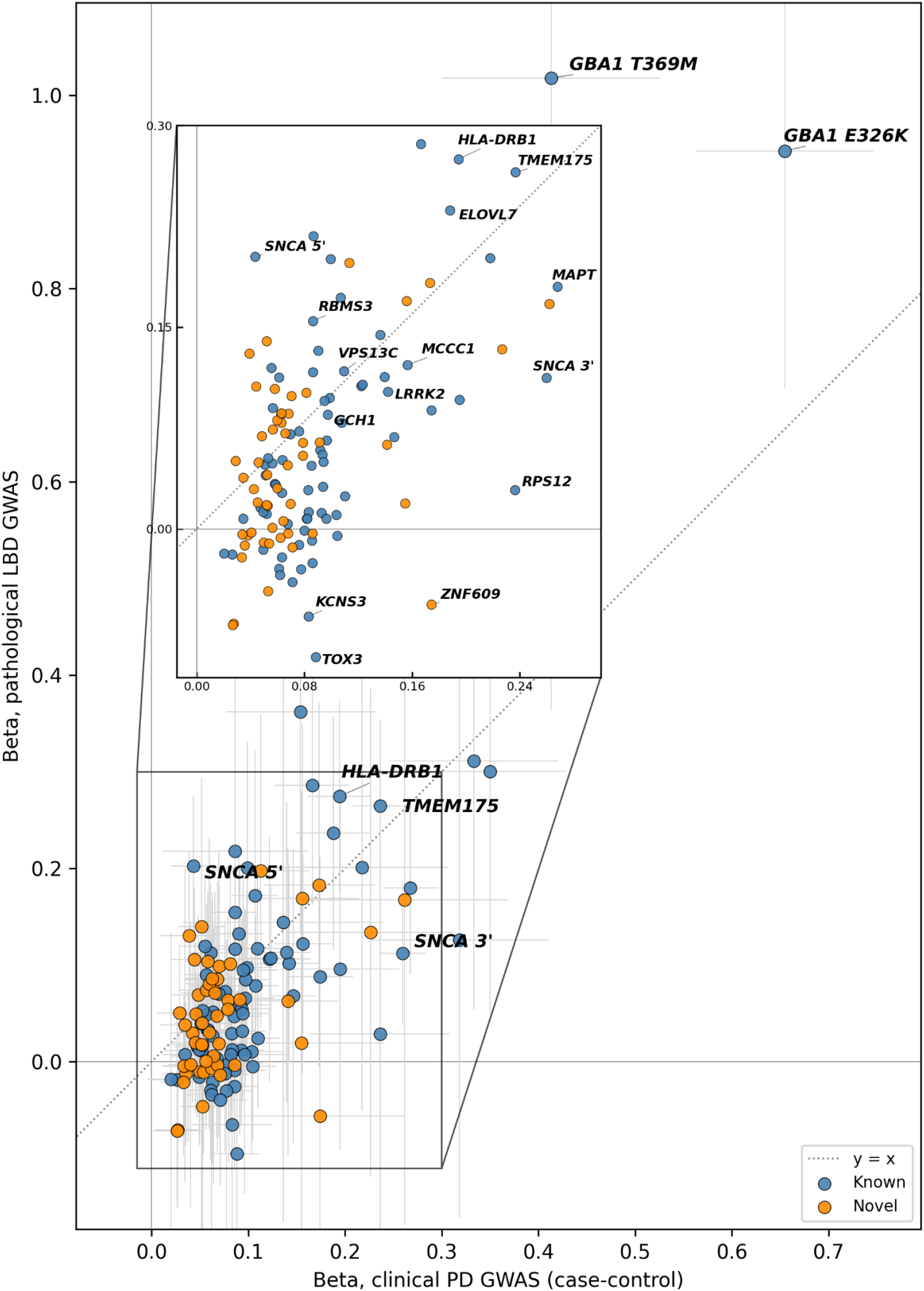
Concordance of clinical Parkinson’s disease and autopsy-confirmed Lewy body disease risk variant effect sizes. Per-allele effect estimates (β) for the 134 out of 157 independent clinical Parkinson’s disease (PD) lead signals that were present and allele-alignable in the pathological Lewy body disease (LBD) GWAS. Each point represents one signal; the x-axis shows the effect in the clinical PD GWAS (European case-control) and the y-axis the effect in the pathological LBD GWAS. All signals are oriented to the PD risk-increasing allele, so PD effects are positive by construction. Points are coloured by their status in the PD GWAS (blue, known; orange, novel); horizontal and vertical grey bars denote 95% confidence intervals. The dotted line marks equality (y = x): points above it have a larger per-allele effect in LBD than in PD, and points below it a smaller one. *SNCA* 5′ and 3′ variants refer to rs2301134 and rs356182, respectively, representing two independent conditional signals at a single locus. Effect sizes were strongly correlated between the two studies (Pearson *r =* 0.78, *P =* 1.8 × 10^−28^), and 103 of 134 signals (77%) were directionally concordant.

Direct comparison of per-allele effect estimates identified a single locus with a significant difference between pathological LBD and clinical PD. At the *SNCA* locus, the 5’ variant rs2301134 showed a larger effect in pathological LBD (b = 0.20, SE = 0.04) than in clinical PD (b = 0.04, SE = 0.01; P difference = 3.75 × 10^−5^; Table 4, Fig. 2), the only comparison among the 134 tested to survive Bonferroni correction. Nominally significant differences were observed at a further 12 variants. At *GBA1* rs2230288 (E326K), the effect was again larger in pathological LBD (b = 0.94, SE = 0.13) than in clinical PD (b = 0.66, SE = 0.05; P difference = 0.03); the remaining 11 had smaller effects in LBD, including *BST1*, *RIT2* and *CRAMP1*, which are genome-wide significant in clinical PD but were not associated here. Other established PD loci, including *LRRK2*, *MAPT*, *GCH1*, *TMEM175* and *VPS13C*, did not differ significantly between studies (all P difference > 0.05; Table 4).

**Table 4.**
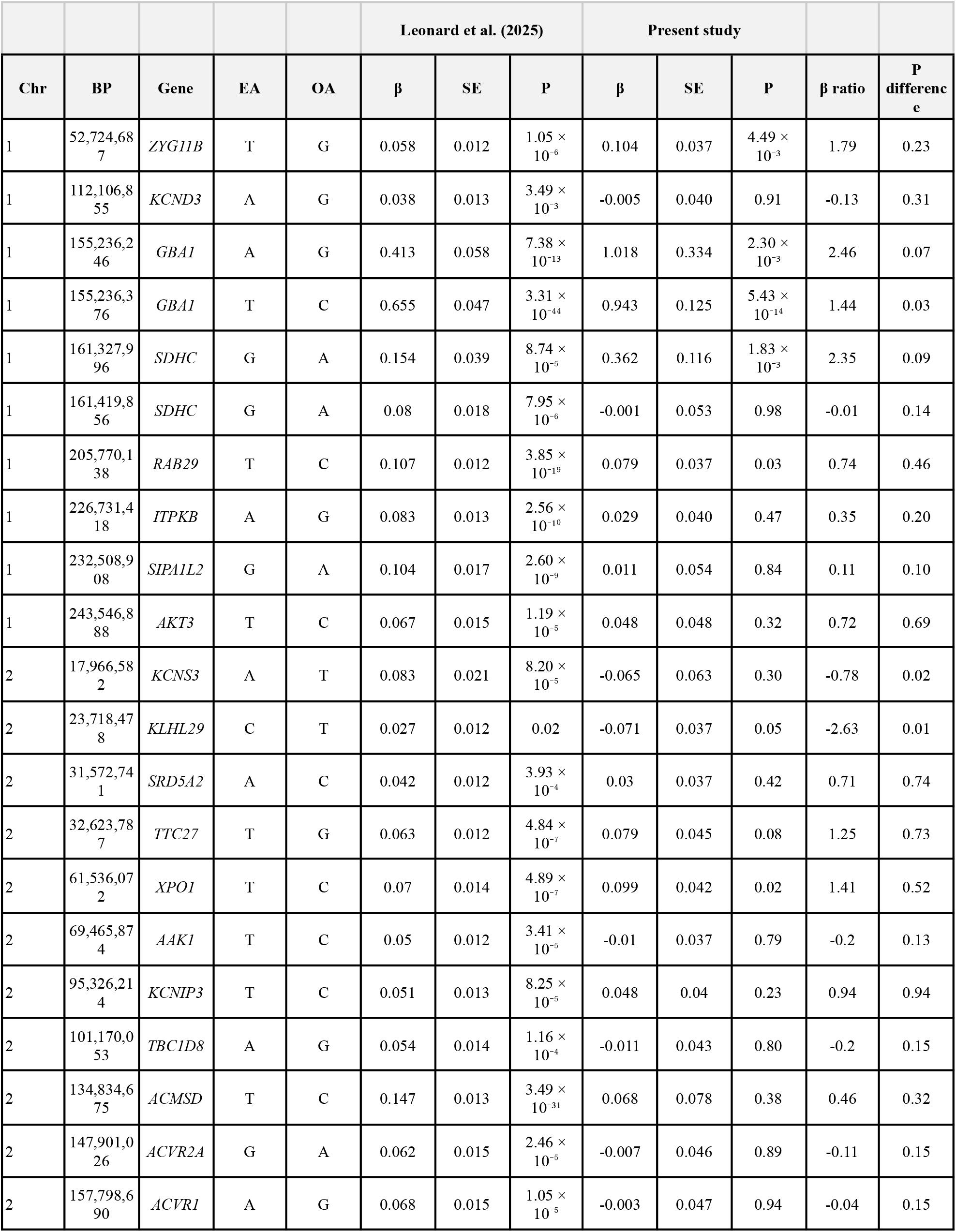
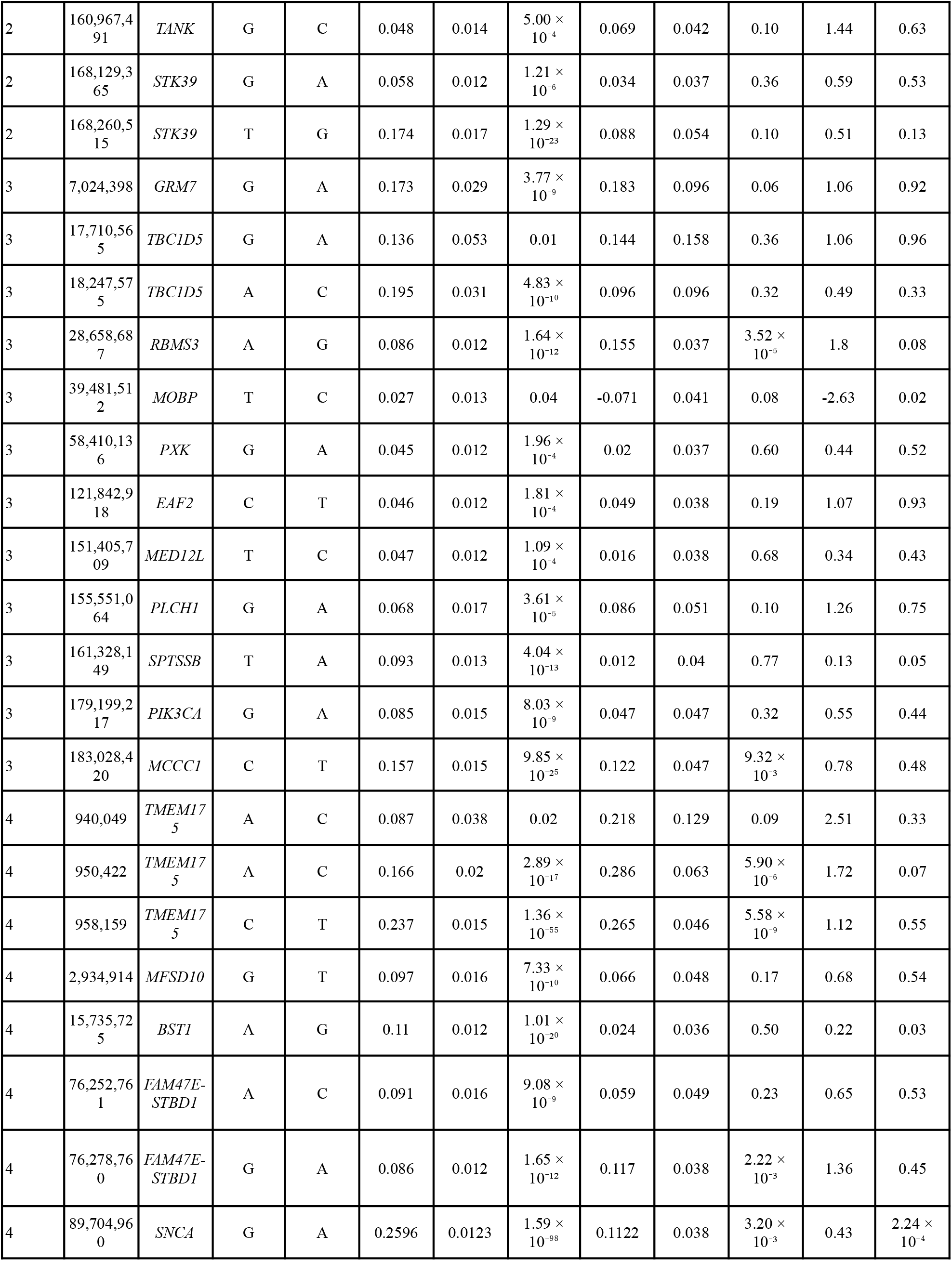
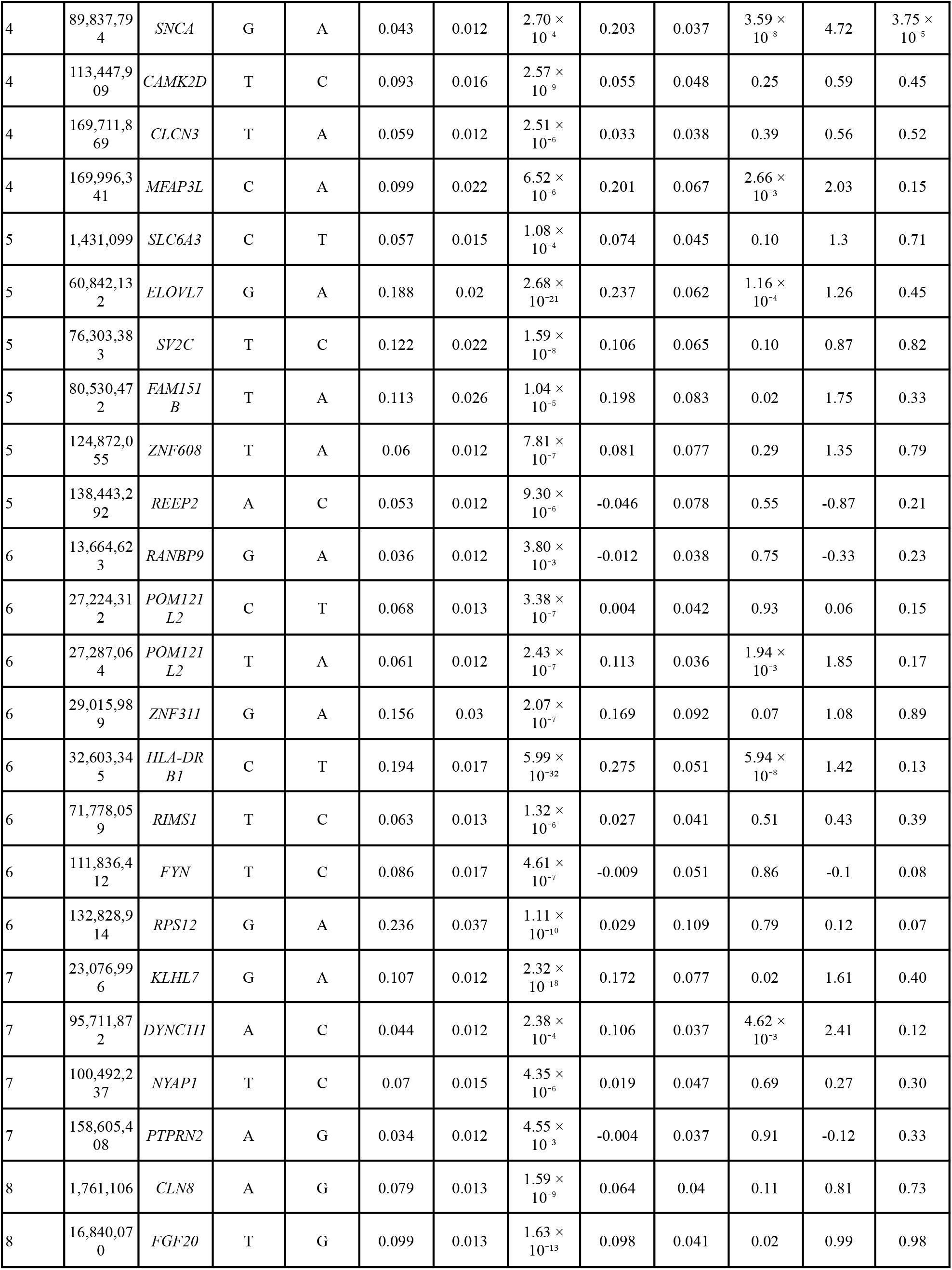
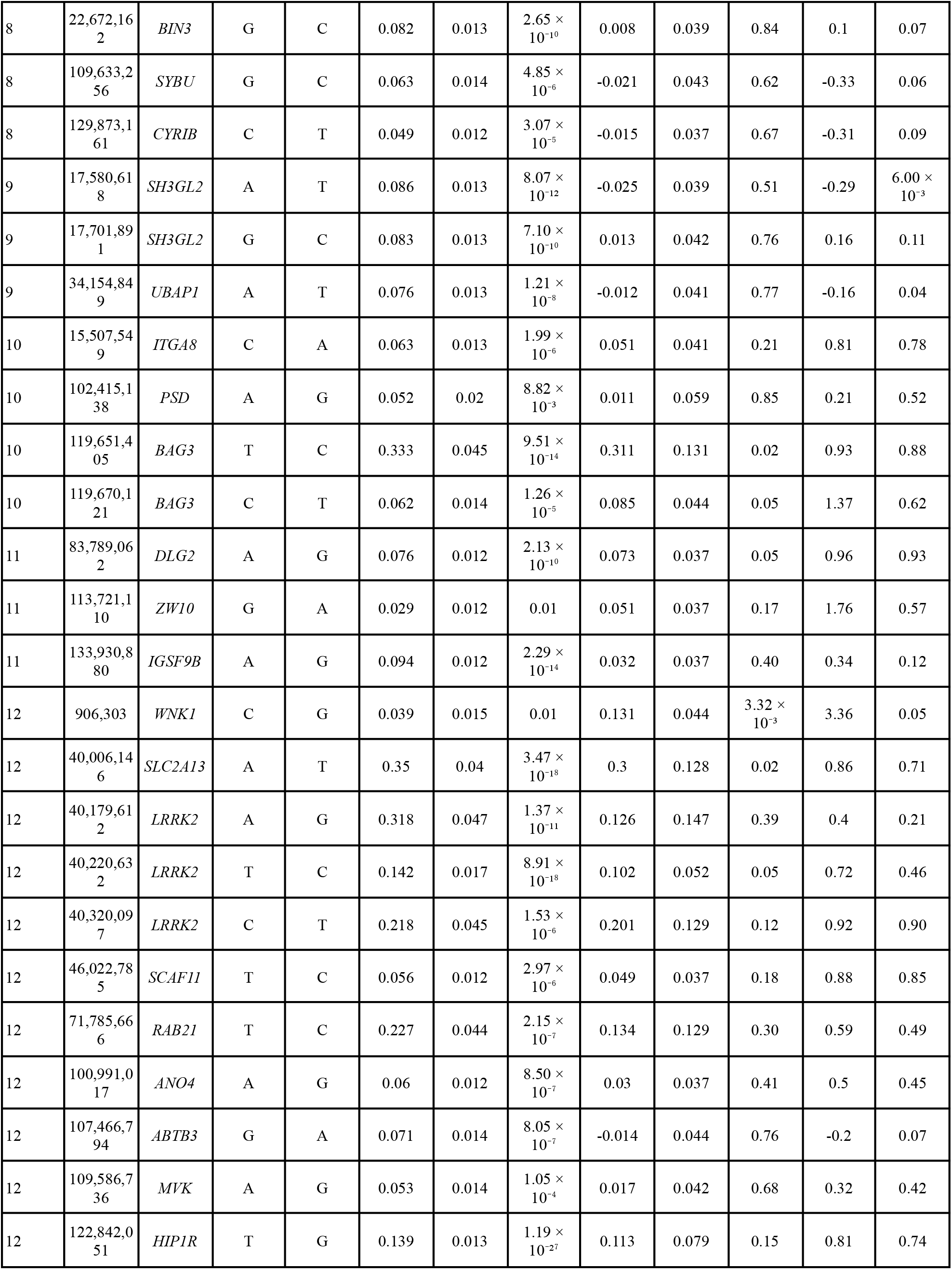
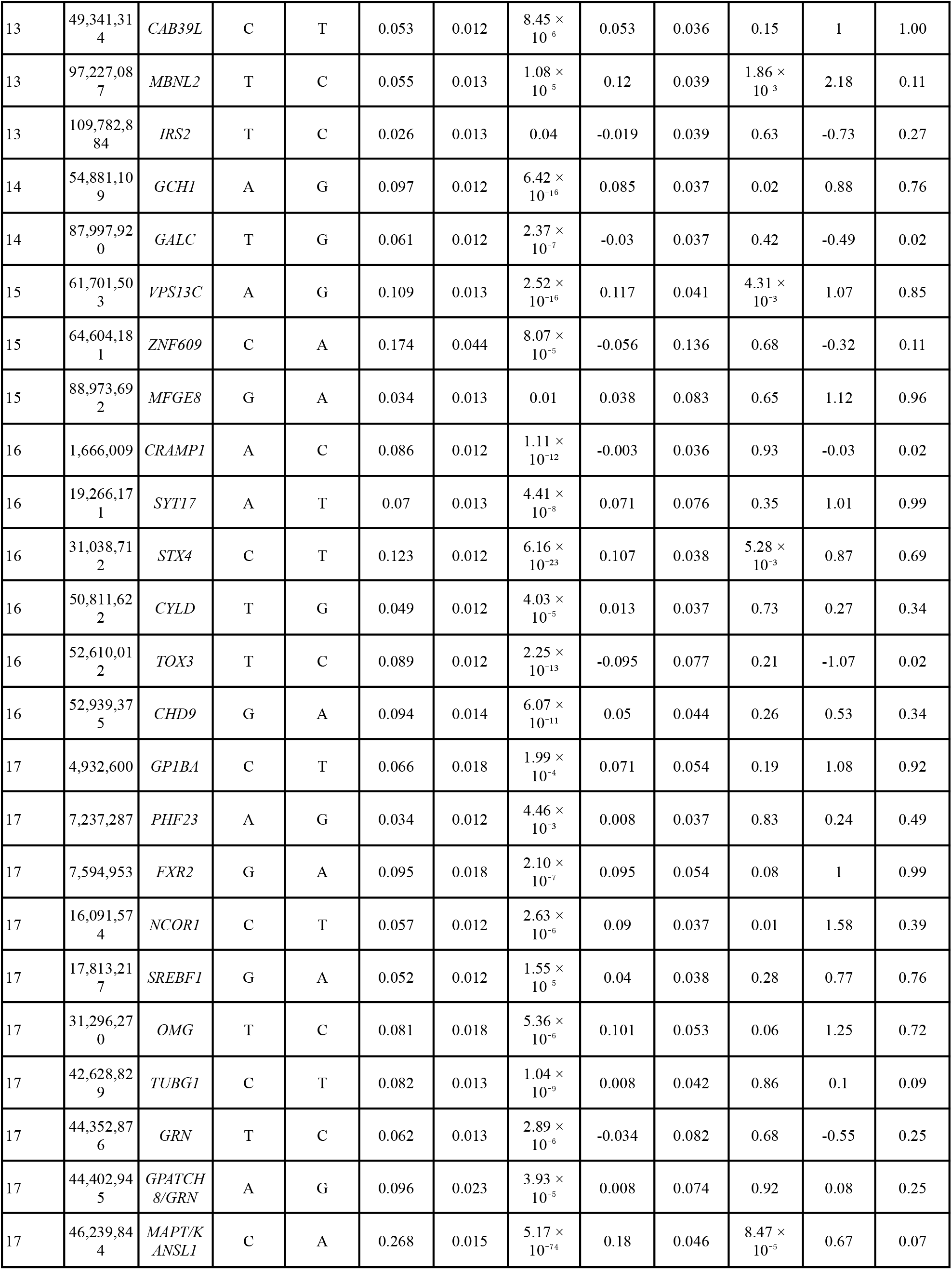
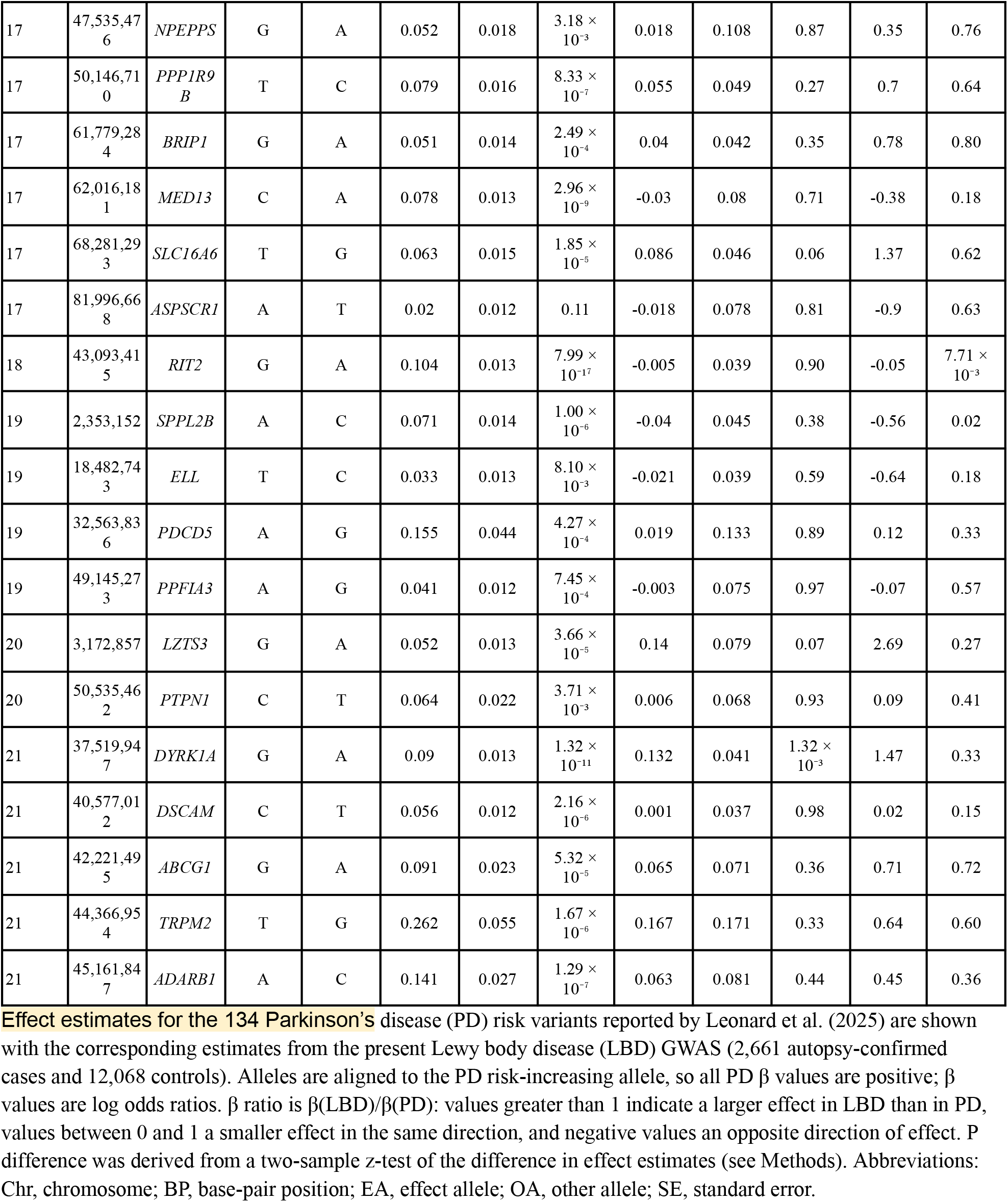
Comparison of effect estimates between clinical PD (Leonard et al., 2025) and the present autopsy-confirmed LBD meta-analysis.

**Table 5.** Cohort characteristics of Lewy body groups across stages.

|  | GP2 |  |  | Mayo |  |  |
| --- | --- | --- | --- | --- | --- | --- |
|  | Brainstem | Limbic | Neocortical | Brainstem | Limbic | Neocortical |
| N | 210 | 257 | 864 | 128 | 352 | 498 |
| Age at death, mean (SD) | 79.6 (8.5) | 79.2 (8.4) | 79.6 (7.4) | 80.2 (10.3) | 80.6 (9.4) | 78.3 (7.9) |
| Sex, F (%) | 77 (36.7) | 80 (31.1) | 298 (34.5) | 52 (40.6) | 150 (42.6) | 212 (42.6) |
| AD co-pathology, n (%) | 113 (53.8) | 78 (30.4) | 474 (54.9) | 68 (53.1) | 269 (76.4) | 416 (83.5) |

In contrast, the genetic correlation with AD was more modest, but remained significant. Using the clinically diagnosed International Genomics of Alzheimer’s Project (IGAP) GWAS ^27^, LBD was positively correlated with AD (rg = 0.33, SE = 0.14, P = 0.02, Table 3). A sensitivity analysis using the larger Bellenguez et al. GWAS, which includes proxy-defined cases, yielded similar estimates (rg = 0.43, SE = 0.13, P = 8.6 × 10^−4^, Table 3), demonstrating that the genetic overlap was consistent between pathological LBD and clinical AD. Given the strong contribution of *APOE* to both AD and our results, we repeated the analyses after excluding the *APOE* region. The genetic correlation between LBD and AD was essentially unchanged (Kunkle rg = 0.31, SE = 0.14, P = 0.03; Bellenguez rg = 0.42, SE = 0.13, P = 0.001), indicating that the shared genetic liability between pathologically confirmed LBD and AD is not driven solely by the *APOE* locus.

### A single genome-wide significant locus distinguishes neocortical from subcortical Lewy body distribution

Having identified five genome-wide significant loci for autopsy-confirmed LBD, we next investigated whether genetic variants influence the anatomical distribution of LB pathology. We performed a case-case genome-wide association analysis comparing individuals with neocortical (n = 1,362) and subcortical (n = 947) LB distribution, comprising 1,331 cases from GP2 and 978 from the Mayo Clinic Brain Bank. Within GP2, 1,331 of 1,928 cases (69.0%) had sufficient Lewy body staging for classification. Significant AD co-pathology was more frequent in neocortical than in subcortical cases in both cohorts (GP2 54.9% versus 40.9%, OR = 1.75, 95% CI 1.39-2.20, P = 1.25 × 10^−06^; Mayo 83.5% versus 70.2%, OR=2.17, 95% CI 1.60-2.95, P = 5.24 × 10^−07^). Neocortical cases also died 2.2 years younger than subcortical cases in Mayo (78.3 versus 80.5 years; 95% CI 1.1-3.3, P = 9.4 × 10^−5^), with no difference in GP2 (79.6 versus 79.4 years, P = 0.59). Analyses were therefore adjusted for AD neuropathologic change, age at death, sex and the first three genetic principal components. Test statistics showed no evidence of residual inflation (λ = 1.006). A single locus reached genome-wide significance, at *APOE*, indexed by the ε4-defining variant rs429358, with the ε4 allele associated with neocortical relative to subcortical distribution (OR = 1.70, 95% CI 1.45-1.98, P = 2.47 × 10^−11^; Figure 1b, Supplementary Figure 4). *APOE* ε4 remained associated with neocortical distribution among cases without significant AD co-pathology (OR = 1.76, 95% CI 1.18-2.63, P = 0.005; 390 neocortical, 276 subcortical), as it was among AD-positive cases (OR = 2.47, 1.70-3.58, P = 2.0 × 10^−6^), with no significant heterogeneity between strata (P = 0.23). Adjustment for age at death and disease duration had little effect on the estimate (Supplementary Table 7).

To determine whether this association extended across the spectrum of LB pathology, we repeated the analysis using an ordinal model of pathological stages (brainstem, limbic, neocortical) in the same dataset. The *APOE*-ε4 risk allele (rs429358) remained the lead signal genome-wide: each ε4 allele was associated with 1.76-fold higher odds of a more widespread LB pathological stage (95% CI 1.50-2.06, P = 1.4 × 10^−12^; Figure 1c). The rs429358 ε4 allele had comparable effects at both stage thresholds (limbic or neocortical vs brainstem-predominant: OR 1.96, 95% CI 1.52-2.52; neocortical vs brainstem-predominant or limbic: OR 1.73, 95% CI 1.47–2.03), with no departure from proportional odds (Brant *P* = 0.51).

The remaining four case-control loci showed no association with Lewy body distribution (Supplementary Table 8). At *TMEM175*, *SNCA* and the *HLA* class II region, power to detect an effect comparable to that at *APOE* exceeded 98% (α = 0.01, corrected for the five loci tested). At the *GBA1* locus, power was 54%, reflecting the low frequency of rs2230288 (2.9%) and heterogeneity between cohorts (I² = 81%).

## Discussion

We performed a genome-wide association meta-analysis of neuropathologically confirmed LBD in 2,661 autopsy-diagnosed cases and 12,068 controls, and identified five genome-wide significant loci: *APOE*, *GBA1*, *SNCA*, *TMEM175* and the *HLA* class II region. Our design addresses two key limitations of clinically defined studies: diagnostic misclassification and the limited ability of clinical syndromes to capture the underlying biological continuum of LB pathology. Defining cases by the presence of LB pathology rather than by antemortem syndrome yielded an architecture that overlapped substantially with clinical PD but was not identical to it. Two features distinguished the pathology-anchored analysis. First, *APOE* was the strongest locus genome-wide. Second, the *SNCA* signal was indexed by a variant distinct from the lead clinical PD variant at the 3′ end of the gene, but identical to the second, 5′ *SNCA* signal reported in DLB and RBD GWAS; importantly, this 5′ signal had a significantly larger effect on LB pathology than the 3′ PD signal. In contrast, many loci established in clinical PD and DLB GWAS did not reach genome-wide significance, which likely reflects, at least in part, differences in statistical power and sample size between studies.

The five associated loci converge on four interconnected biological processes: lysosomal function (*GBA1, TMEM175*), α-synuclein expression (*SNCA*), adaptive immunity (*HLA* class II), and lipid transport and glial clearance (*APOE*). Three of these intersect with the handling and clearance of α-synuclein rather than its synthesis. *GBA1* and *TMEM175* influence lysosomal degradation of α-synuclein ^33,34^; *HLA* class II molecules bind and traffic abnormal α-synuclein extracellularly ^35^, with recent experimental work in mice implicating specific alleles in its aggregation ^36^; and *APOE*ε4-expressing astrocytes showed reduced re-uptake of α-synuclein and impaired autophagy *in vitro* ^37^. The two largest effects in our study (*GBA1*, OR = 2.57, 95% CI 2.01-3.28; *APOE*, OR = 2.27, 95% CI 2.06-2.50) arise at clearance-associated loci. *SNCA, TMEM175* and (more weakly) *GBA1*, have also been associated with age at onset in clinical PD ^38^, implicating these loci in the timing of disease as well as its pathology. Together, these convergences point to the capacity to process and clear α-synuclein as a candidate determinant of LB pathology, and one that an autopsy-confirmed endpoint is particularly suited to resolve.

*APOE* was the only locus to reach genome-wide significance in the case-only analysis, with the ε4 allele associated with neocortical relative to subcortical LB pathology after adjustment for concomitant AD co-pathology and age at death. Age at death was included because pathology accumulates with age and because ε4 is itself associated with age at onset (Voss et al., in press), with mortality in PD, ^39^, and with lifespan in the general population independently of neurodegenerative disease ^40^; the estimate was not attenuated when it was omitted. The association was likewise not explained by measured AD co-pathology: ε4 was associated with neocortical distribution in cases without significant AD co-pathology as well as in those with it, with no significant difference between strata. Thal phase and Braak neurofibrillary stage are nonetheless coarse ordinal summaries, and cases classified as lacking significant AD co-pathology are defined by a threshold rather than by the absence of amyloid or tau, so residual confounding by unmeasured variation in amyloid or tau burden cannot be excluded. A recent GWAS of neuropathological outcomes in LBD cases likewise associated ε4 with a more severe LBD subtype, which attenuated on adjustment for Braak stage and Thal phase overall but persisted in cases with low AD pathology ^41^. However, that study used cases from the Mayo Clinic Jacksonville Brain Bank, which contributes 978 of the 2,309 cases analysed here; the two analyses are therefore not independent. Nevertheless, the non-overlapping GP2 cohort provides independent support for this association. A mechanism independent of amyloid and tau is further supported by experimental work demonstrating that *APOEε4* exacerbates α-synuclein pathology in transgenic mice in the absence of Aβ deposition ^42,43^. Together with the case-control association reported above, these findings support a dual role for *APOE*, influencing both the likelihood of developing LB pathology and its anatomical distribution through mechanisms at least partly independent of amyloid and tau.

The *HLA* class II region is an established susceptibility locus in clinical PD, and its recovery under pathological ascertainment shows that the association is also present when cases are defined by LB pathology. Le Guen *et al*. (2023) observed only a subthreshold protective association at this locus in the previous DLB GWAS (OR = 0.91, P = 0.07), which they attributed to limited power ^44^. In our data, *HLA-DQB1\**03:02 and *HLA-DQA1\**03:01 were both associated with reduced risk (OR = 0.72, 95% CI 0.59-0.87, and OR = 0.71, 95% CI 0.58-0.86, respectively). Both alleles are carried on haplotypes containing *DRB1*\*04 subtypes, and previous fine-mapping of the *HLA* locus in AD and PD has suggested that the association is primarily driven by *DRB1*\*04 rather than the linked *DQA1*\*03:01 / *DQB1*\*03:02 alleles, since *DRB1*\*04 haplotypes carrying other DQ alleles remain associated whereas *DQA1*\*03:01 / *DQB1*\*03:02 on a *DRB1*\*08:02 background does not ^44^. Our concordant protective estimate for *DRB1*\*04:04, the subtype with the strongest reported effect, is consistent with this interpretation.

An informative absence from our results is *BIN1*, which reached genome-wide significance in the DLB GWAS by Chia et al. ^1^ but showed no association in our cohorts (OR = 1.01, 95% CI 0.94-1.09, P = 0.76). Our estimate is significantly lower than theirs (nominal P difference = 0.03), suggesting the absence reflects more than limited power. Given that the *BIN1* association has been linked more closely to neurofibrillary tangle burden than to LB pathology ^45^, a dementia-based case definition may enrich tangle-associated loci relative to ours. At the 17q21 (*MAPT* locus), the clinical PD index variant was associated with autopsy-confirmed LBD at the multiple-testing threshold applied across the 134 PD lead variants examined, although it did not reach genome-wide significance in the primary analysis. Its per-allele effect was lower than that reported in clinical PD (β ratio = 0.67, P difference = 0.07). Although this difference was not significant, two non-exclusive explanations could account for a smaller effect under pathological ascertainment. First, clinically ascertained PD series may include individuals with progressive supranuclear palsy (PSP) or corticobasal degeneration, for which the H1 *MAPT* haplotype is the principal genetic risk factor ^46,47^. Second, part of the PD risk at this locus may act through mechanisms not directly tied to LB pathology: the H1 *MAPT* haplotype has been associated with reduced expression of *KANSL1*, a regulator of *PINK1*-dependent mitophagy ^48^, and *PRKN*-associated parkinsonism, which involves the same pathway, frequently occur without LB pathology ^49^. A more limited contribution of *MAPT* to LB pathology specifically is also consistent with the wider literature: *MAPT* was not identified in the largest Lewy body dementia sequencing study ^1^ or an independent European DLB GWAS ^50^, while in autopsy-confirmed cases, common H1/H2 haplotypes showed no association with Lewy body burden, with only a rare H1 subhaplotype surviving correction for multiple testing ^51^. MAPT has likewise not been associated with age at onset ^38^, further suggesting that its contribution to PD risk does not extend to the Lewy body phenotype.

Our study has several limitations. Autopsy ascertainment defines our case series but also selects it: brain donors differ systematically from the general population, and variants influencing survival to post-mortem may be differentially represented among cases and controls, and preferential recruitment of donors with rapidly progressive disease or prominent dementia would compound this selection, which may contribute to the differences we observe relative to syndrome-anchored studies. Neuropathological assessment was semi-quantitative and conducted across multiple centres over several decades, during which different staging schemes were in use, and regional sampling and the application of staging criteria, which involve an element of subjective judgement, are likely to have varied between centres and neuropathologists. AD co-pathology was common among cases and because it may both confound and mediate associations at loci such as *APOE*, its contribution cannot be fully separated from that of LB pathology. Our analyses were restricted to individuals of European ancestry, reflecting the composition of the contributing brain banks; extending autopsy-confirmed collections beyond European-ancestry donors is important both for equity and because differing patterns of linkage disequilibrium could improve fine-mapping resolution at these loci. Finally, although the pathology-anchored design is a principal strength of this study, it constrains sample size and provides a single cross-sectional assessment of pathology at death, which may reflect any stage of disease depending on disease duration and competing causes of death, and precludes direct inference about progression. Seed amplification assays (SAA) detect pathological α-synuclein in cerebrospinal fluid with high accuracy and have been validated against post-mortem examination, offering the prospect of a molecularly defined case definition in living cohorts. Sensitivity may be lower when LB pathology is limited or brainstem-predominant, and seeding activity varies with genetic status, meaning that such cohorts would introduce ascertainment biases of their own. Nevertheless, they could enable substantially larger case series than brain banking permits while retaining a biologically anchored case definition, and provide an opportunity to test the loci identified here in SAA-positive and SAA-negative PD.

Overall, by defining cases by LB pathology rather than by clinical syndrome, we identified five loci associated with Lewy body disease, implicating lysosomal function, α-synuclein biology, adaptive immunity and lipid metabolism. *APOE* was the strongest locus genome-wide and was further associated with neocortical rather than subcortical distribution of LB pathology, indicating a role in both the development of LB pathology and its anatomical extent. Integrating neuropathological and genetic data in this way identifies determinants of the anatomical extent of LB that syndrome-anchored studies cannot resolve, and provides a basis for prioritising therapeutic targets in Lewy body disease.

## Contributors

LYW analysed the data and drafted the manuscript. JT contributed to the HLA data analysis. HRM conceptualised and supervised the study.

## Declaration of Interest

HRM received grant funding and support for attending meetings or travel from the Michael J Fox Foundation for Parkinson’s Research for this work; received grant funding from the Progressive Supranuclear Palsy Association, Corticobasal Disease Solutions, Drake Foundation, Parkinson’s UK, Medical Research Council UK, and Cure Parkinson’s Trust unrelated to this work; received consulting fees from Roche, Amylyx, and Aprinoia; received honoraria from Kyowa-Kirin, *British Medical Journal*, and the Movement Disorders Society; is a coapplicant on a patent application related to a C9ORF72 method for diagnosing a neurodegenerative disease; and serves on the Cure Progressive Supranuclear Palsy Association Advisory Board, the Association of British Neurologists Movement Disorders Special Interest Group, and the Association of British Neurologists Neurogenetics Advisory Group.

## Data and code availability

Data used in the preparation of this article were obtained from the Global Parkinson’s Genetics Program (GP2; https://gp2.org). Specifically, we used Tier 2 data from GP2 release 11 (https://zenodo.org/records/17753486). Clinical, demographic, and neuropathological data for NIH NeuroBioBank donors were obtained from the NeuroBioBank Portal (https://neurobiobank.nih.gov/). Whole-genome sequencing data for these donors were obtained through a data access application to the NIMH Data Archive (collection #3917; https://nda.nih.gov/nbb) Clinical, neuropathological, and genetic data from the Mayo Clinic Jacksonville Brain Bank were obtained on request from the brain bank. See Supplementary Table 9 for key resources used in this project All code generated for this article, and the identifiers for all software programs and packages used, are available on GitHub.

## Acknowledgement

We would like to thank all the donors and their families for their invaluable contribution, as well as the collaborating brain banks for their essential support. This research was funded by the UKRI/MRC award - Defining and diagnosing neurodegenerative Movement Disorders through integrated analysis of Genetics And neuroPathology (MD-GAP) (MR/T018569/1), and the Global Parkinson’s Genetic Program (GP2). This project was supported by the Global Parkinson’s Genetics Program (GP2; https://gp2.org), a research program of Aligning Science Across Parkinson’s (ASAP). ASAP is managed by the Coalition for Aligning Science (CAS) (https://ror.org/00rf4ez40). The Michael J. Fox Foundation for Parkinson’s Research (MJFF) (https://ror.org/03arq3225) is the funding and strategic partner. For a complete list of GP2 members see https://doi.org/10.5281/zenodo.7904831. This research was funded in part by Aligning Science Across Parkinson’s ASAP-000478 through the Michael J. Fox Foundation for Parkinson’s Research (MJFF). This paper presents independent research supported by the NIHR, Newcastle Biomedical Research Centre (BRC). Funding for Collection 3917, Whole-Genome Sequence Analysis of Postmortem Human Brains from the NIH NeuroBioBank, in the NIMH Data Archive (https://nda.nih.gov/) was provided by the National Institute of Mental Health. The Arizona Study of Aging and Neurodegenerative Disorders and Brain and Body Donation Program at Banner Sun Health Research Institute has been supported by the National Institute of Neurological Disorders and Stroke (U24 NS072026 National Brain and Tissue Resource for Parkinson’s Disease and Related Disorders), the National Institute on Aging (P30 AG019610 and P30AG072980, Arizona Alzheimer’s Disease Center), the Arizona Department of Health Services (contract 211002, Arizona Alzheimer’s Research Center), the Arizona Biomedical Research Commission (contracts 4001, 0011, 05-901 and 1001 to the Arizona Parkinson’s Disease Consortium) and the Michael J. Fox Foundation for Parkinson’s Research. Tissue samples were supplied by The Manchester Brain Bank, which is part of the Brains for Dementia Research programme, jointly funded by Alzheimer’s Research UK and Alzheimer’s Society. This research was supported in part by the Intramural Research Program of the National Institutes of Health (NIH; program #: ZIAAG000935, ZIANS003154). The contributions of the NIH authors were made as part of their official duties as NIH federal employees, are in compliance with agency policy requirements, and are considered works of the United States Government. However, the findings and conclusions presented in this paper are those of the authors and do not necessarily reflect the views of the NIH or the U.S. Department of Health and Human Services. For the purpose of open access, the author has applied a CC BY public copyright licence to all Author Accepted Manuscripts arising from this submission.

## Notes

### Author Declarations

The present study was approved by the University College London Research Ethics Committee (23473/001).

