## Supplementary Material for "Genetic susceptibility to Lewy body disease and the distribution of Lewy pathology"

#### Supplementary Table 1. Equivalence between Lewy body pathology staging systems.

| Unified staging system for LBD | McKeith stage | Lewy Body Braak stage |
|---|---|---|
| Neocortical | Neocortical | Stages 5–6 |
| Limbic | Limbic | Stage 4 |
| Brainstem | Brainstem | Stages 1–3 |

A table showing equivalence between different staging systems. Based on BrainNet Europe Protocol.

#### Supplementary Table 2. Derivation of Alzheimer's disease neuropathologic change score from Thal Aβ-phase and Braak NFT stage.

| | Braak NFT 0 | Braak NFT I–II | Braak NFT III–IV | Braak NFT V–VI |
|---|---|---|---|---|
| Thal Aβ-phase 0 | No | No | No | No |
| Thal Aβ-phase 1–2 | No | Low | Low | Low |
| Thal Aβ-phase 3 | No | Low | Intermediate | Intermediate |
| Thal Aβ-phase 4–5 | No | Low | Intermediate | High |

An adaptation of the National Institute on Aging and the Alzheimer’s Association (NIA-AA) criteria, in which Thal phases and Braak NFT were integrated to generate an AD score. From Tunold et al. (2023).

#### Supplementary Table 3. Demographic and clinicopathological characteristics of the PD/PDD only and controls GWAS.

| Clinical Diagnosis | PD | PDD | Controls |
|---|---|---|---|
| Pathological Diagnosis | LBD | LBD | - |
| N | 890 | 326 | 10517 |
| Sex (F, %) | 342 (38.4%) | 94 (28.8%) | 5577 (53.0%) |
| Age (mean, sd) | 78.8 (7.6) | 77.8 (6.5) | 63.4 (14.0) |

Only includes individuals from GP2.

#### Supplementary Table 4. Lead SNPs identified in the PD/PDD-only versus control GWAS.

| | | | | | | PD/PDD only | | All GP2 LBD cases | |
|---|---|---|---|---|---|---|---|---|---|
| rsID | Chr | BP | A1 | EAF | Nearest gene | OR (95% CI) | P | OR (95% CI) | P |
| rs2230288 | 1 | 155,236,376 | T | 0.014 | *GBA1* | 2.80 (2.07–3.77) | $1.46 \times 10^{-11}$ | 2.38 (1.81–3.13) | $5.64 \times 10^{-10}$ |
| rs34311866 | 4 | 958,159 | C | 0.182 | *TMEM175* | 1.25 (1.11–1.41) | $2.0 \times 10^{-4}$ | 1.26 (1.14–1.40) | $8.30 \times 10^{-6}$ |
| rs356182 | 4 | 89,704,960 | G | 0.346 | *SNCA* | 1.35 (1.22–1.49) | $2.74 \times 10^{-9}$ | 1.17 (1.07–1.27) | $3.22 \times 10^{-4}$ |
| rs2737030 | 4 | 89,788,948 | T | 0.789 | *SNCA* | 1.29 (1.14–1.47) | $5.06 \times 10^{-5}$ | 1.39 (1.24–1.54) | $3.37 \times 10^{-9}$ |
| rs3117443 | 6 | 29,384,854 | A | 0.591 | extended MHC class I | 0.74 (0.67–0.81) | $7.81 \times 10^{-10}$ | 0.81 (0.75–0.88) | $7.03 \times 10^{-7}$ |
| rs9275382 | 6 | 32,701,054 | T | 0.105 | *HLA class II* | 0.68 (0.57–0.80) | $3.14 \times 10^{-6}$ | 0.65 (0.56–0.75) | $4.87 \times 10^{-9}$ |
| rs429358 | 19 | 44,908,684 | C | 0.126 | *APOE* | 1.57 (1.37–1.81) | $1.20 \times 10^{-10}$ | 2.18 (1.95–2.44) | $2.38 \times 10^{-43}$ |

Association statistics for lead variants in the PD/PDD-only case–control analysis, with the corresponding estimates in all GP2 Lewy body disease cases.

Variants comprise the five loci identified in the primary meta-analysis (Table 2) together with the two additional lead variants identified in the PD/PDD-only analysis, rs356182 and rs3117443.

Odds ratios are expressed per copy of the effect allele (A1) and are oriented identically in the two analyses.

Positions are in GRCh38.

Genome-wide significance was defined as $P < 5 \times 10^{-8}$.

**Supplementary Table 5. Association of classical HLA alleles with neuropathologically confirmed Lewy body disease.**

| Allele | Cases, n (%) | Controls, n (%) | OR (95% CI) | P | q |
|---|---|---|---|---|---|
| HLA-DQB1*03:02 | 164 (16.0%) | 523 (20.6%) | 0.72 (0.59–0.87) | $5.40 \times 10^{-4}$ | 0.0201 |
| HLA-DQA1*03:01 | 149 (14.9%) | 476 (19.3%) | 0.71 (0.58–0.86) | $5.59 \times 10^{-4}$ | 0.0201 |
| HLA-C*05:01 | 215 (21.5%) | 419 (16.9%) | 1.27 (1.05–1.53) | 0.0125 | 0.231 |
| HLA-A*01:01 | 354 (34.6%) | 740 (29.0%) | 1.19 (1.04–1.37) | 0.0128 | 0.231 |
| HLA-B*44:02 | 211 (20.6%) | 421 (16.7%) | 1.25 (1.04–1.51) | 0.0174 | 0.233 |
| HLA-DRB1*07:01 | 286 (29.0%) | 632 (25.4%) | 1.21 (1.03–1.42) | 0.0199 | 0.233 |
| HLA-DQB1*02:02 | 221 (21.7%) | 475 (18.7%) | 1.23 (1.03–1.46) | 0.0227 | 0.233 |
| HLA-A*03:01 | 266 (25.7%) | 765 (29.4%) | 0.83 (0.71–0.98) | 0.0263 | 0.237 |
| HLA-C*08:02 | 82 (8.3%) | 163 (6.6%) | 1.33 (0.99–1.77) | 0.0544 | 0.375 |
| HLA-C*02:02 | 74 (7.5%) | 225 (9.1%) | 0.77 (0.58–1.01) | 0.0623 | 0.375 |
| HLA-B*08:01 | 264 (26.0%) | 544 (21.5%) | 1.18 (0.99–1.39) | 0.0624 | 0.375 |
| HLA-DRB1*03:01 | 276 (27.5%) | 588 (23.5%) | 1.17 (0.99–1.39) | 0.0687 | 0.375 |
| HLA-DQB1*02:01 | 273 (26.8%) | 587 (23.2%) | 1.15 (0.99–1.35) | 0.0735 | 0.375 |
| HLA-DQA1*05:01 | 276 (27.4%) | 589 (23.8%) | 1.16 (0.99–1.36) | 0.0736 | 0.375 |
| HLA-DQA1*05:05 | 204 (20.1%) | 600 (24.0%) | 0.85 (0.70–1.02) | 0.0782 | 0.375 |
| HLA-DRB1*04:04 | 66 (6.7%) | 208 (8.4%) | 0.78 (0.58–1.04) | 0.0901 | 0.405 |
| HLA-B*35:01 | 96 (9.5%) | 297 (11.8%) | 0.81 (0.64–1.04) | 0.0980 | 0.415 |
| HLA-DQA1*02:01 | 287 (28.7%) | 636 (25.8%) | 1.14 (0.97–1.33) | 0.110 | 0.440 |
| HLA-A*68:01 | 55 (5.4%) | 176 (6.9%) | 0.78 (0.56–1.07) | 0.124 | 0.470 |
| HLA-C*03:03 | 132 (13.4%) | 285 (11.5%) | 1.18 (0.95–1.48) | 0.135 | 0.486 |
| HLA-B*40:01 | 94 (9.3%) | 257 (10.2%) | 0.84 (0.65–1.09) | 0.185 | 0.575 |
| HLA-B*13:02 | 43 (4.3%) | 151 (6.0%) | 0.79 (0.55–1.12) | 0.185 | 0.575 |
| HLA-DQB1*06:04 | 53 (5.2%) | 181 (7.2%) | 0.80 (0.58–1.11) | 0.186 | 0.575 |
| HLA-B*44:03 | 114 (11.3%) | 253 (10.1%) | 1.17 (0.92–1.49) | 0.192 | 0.575 |
| HLA-DRB1*13:02 | 75 (7.6%) | 234 (9.5%) | 0.83 (0.63–1.11) | 0.207 | 0.597 |
| HLA-DPB1*04:02 | 219 (24.7%) | 510 (23.0%) | 1.12 (0.93–1.35) | 0.225 | 0.624 |
| HLA-DQB1*03:03 | 87 (8.5%) | 235 (9.3%) | 0.87 (0.67–1.13) | 0.289 | 0.770 |
| HLA-C*12:03 | 72 (7.3%) | 230 (9.3%) | 0.87 (0.65–1.15) | 0.323 | 0.824 |
| HLA-DQA1*01:02 | 380 (37.2%) | 983 (38.6%) | 0.93 (0.80–1.08) | 0.347 | 0.824 |
| HLA-DPB1*01:01 | 124 (14.2%) | 262 (12.0%) | 1.11 (0.88–1.40) | 0.380 | 0.824 |
| HLA-C*16:01 | 85 (8.6%) | 191 (7.7%) | 1.13 (0.86–1.47) | 0.381 | 0.824 |
| HLA-DPB1*02:01 | 247 (28.0%) | 669 (30.2%) | 0.93 (0.78–1.10) | 0.387 | 0.824 |
| HLA-C*06:02 | 173 (17.5%) | 472 (19.0%) | 0.92 (0.76–1.11) | 0.389 | 0.824 |
| HLA-B*14:02 | 57 (5.6%) | 129 (5.1%) | 1.15 (0.83–1.59) | 0.389 | 0.824 |
| HLA-B*57:01 | 65 (6.4%) | 171 (6.8%) | 0.89 (0.66–1.20) | 0.430 | 0.845 |
| HLA-DQA1*03:03 | 167 (16.7%) | 363 (14.7%) | 1.08 (0.89–1.31) | 0.447 | 0.845 |

| | | | | | |
|---|---|---|---|---|---|
| HLA-A*32:01 | 86 (8.4%) | 198 (7.8%) | 1.11 (0.85–1.45) | 0.448 | 0.845 |
| HLA-A*24:02 | 180 (17.5%) | 431 (16.9%) | 1.07 (0.89–1.30) | 0.458 | 0.845 |
| HLA-B*15:01 | 141 (13.9%) | 313 (12.5%) | 1.08 (0.87–1.34) | 0.497 | 0.845 |
| HLA-DQB1*05:01 | 245 (24.0%) | 584 (22.9%) | 1.06 (0.90–1.25) | 0.506 | 0.845 |
| HLA-C*03:04 | 145 (14.7%) | 363 (14.6%) | 0.93 (0.76–1.15) | 0.508 | 0.845 |
| HLA-A*29:02 | 79 (7.7%) | 178 (7.0%) | 1.10 (0.84–1.44) | 0.508 | 0.845 |
| HLA-A*11:01 | 123 (12.0%) | 293 (11.5%) | 1.08 (0.86–1.34) | 0.516 | 0.845 |
| HLA-DQA1*01:01 | 228 (22.7%) | 552 (22.2%) | 1.06 (0.89–1.26) | 0.517 | 0.845 |
| HLA-DRB1*11:01 | 100 (10.1%) | 290 (11.7%) | 0.93 (0.72–1.19) | 0.544 | 0.858 |
| HLA-DQB1*04:02 | 55 (5.4%) | 126 (5.0%) | 1.10 (0.79–1.54) | 0.560 | 0.858 |
| HLA-C*07:01 | 317 (31.6%) | 740 (29.5%) | 1.05 (0.90–1.23) | 0.560 | 0.858 |
| HLA-DQB1*06:03 | 132 (13.0%) | 328 (13.0%) | 1.06 (0.85–1.32) | 0.599 | 0.893 |
| HLA-B*27:05 | 76 (7.5%) | 185 (7.4%) | 0.93 (0.70–1.23) | 0.607 | 0.893 |
| HLA-B*51:01 | 88 (8.7%) | 263 (10.4%) | 0.94 (0.72–1.23) | 0.658 | 0.948 |
| HLA-DQA1*01:03 | 136 (13.6%) | 347 (14.0%) | 1.05 (0.85–1.29) | 0.679 | 0.950 |
| HLA-DRB1*01:01 | 183 (18.5%) | 446 (18.0%) | 1.04 (0.86–1.27) | 0.688 | 0.950 |
| HLA-DRB1*04:07 | 58 (5.9%) | 127 (5.2%) | 1.07 (0.77–1.48) | 0.705 | 0.950 |
| HLA-DRB1*14:54 | 59 (6.0%) | 142 (5.8%) | 1.06 (0.77–1.47) | 0.713 | 0.950 |
| HLA-C*04:01 | 181 (18.2%) | 513 (20.6%) | 0.97 (0.80–1.17) | 0.730 | 0.955 |
| HLA-DQB1*06:02 | 289 (28.3%) | 683 (27.0%) | 0.98 (0.84–1.14) | 0.791 | 0.992 |
| HLA-B*07:02 | 293 (28.8%) | 695 (27.3%) | 0.98 (0.84–1.15) | 0.792 | 0.992 |
| HLA-C*07:02 | 312 (31.4%) | 744 (29.7%) | 0.98 (0.84–1.14) | 0.799 | 0.992 |
| HLA-DRB1*04:01 | 181 (18.4%) | 410 (16.6%) | 0.98 (0.80–1.19) | 0.833 | 0.996 |
| HLA-DPB1*04:01 | 712 (69.7%) | 1745 (68.9%) | 1.01 (0.88–1.18) | 0.846 | 0.996 |
| HLA-DRB1*13:01 | 127 (12.9%) | 317 (12.9%) | 1.02 (0.81–1.28) | 0.881 | 0.996 |
| HLA-DPB1*03:01 | 198 (22.7%) | 470 (21.5%) | 0.99 (0.82–1.19) | 0.905 | 0.996 |
| HLA-C*01:02 | 75 (7.6%) | 168 (6.8%) | 0.99 (0.74–1.32) | 0.931 | 0.996 |
| HLA-DQA1*01:04 | 62 (6.2%) | 166 (6.7%) | 0.99 (0.72–1.35) | 0.937 | 0.996 |
| HLA-A*02:01 | 513 (49.8%) | 1227 (47.8%) | 1.00 (0.88–1.13) | 0.962 | 0.996 |
| HLA-B*18:01 | 90 (8.9%) | 236 (9.4%) | 1.00 (0.77–1.30) | 0.978 | 0.996 |
| HLA-DPB1*05:01 | 47 (5.4%) | 119 (5.5%) | 1.00 (0.71–1.41) | 0.981 | 0.996 |
| HLA-DPB1*11:01 | 46 (5.3%) | 115 (5.3%) | 1.00 (0.70–1.44) | 0.984 | 0.996 |
| HLA-A*26:01 | 52 (5.1%) | 145 (5.7%) | 1.00 (0.72–1.38) | 0.985 | 0.996 |
| HLA-DQB1*03:01 | 341 (32.8%) | 866 (33.6%) | 1.00 (0.86–1.16) | 0.987 | 0.996 |
| HLA-DQB1*05:03 | 62 (6.1%) | 169 (6.7%) | 1.00 (0.73–1.37) | 0.993 | 0.996 |
| HLA-DRB1*15:01 | 297 (29.7%) | 700 (27.9%) | 1.00 (0.85–1.18) | 0.996 | 0.996 |

q: BH across the 72 pre-specified classical alleles, within each encoding
Alleles are ordered by P.

**Supplementary Table 6. Genome-wide gene-based association results (MAGMA) for the autopsy-confirmed Lewy body disease case-control GWAS meta-analysis.**

| Gene symbol | Ensembl gene ID | Chr | Start (GR37) | Stop (GR37) | SNPs (n) | Z | P | P (Bonferroni) |
|---|---|---|---|---|---|---|---|---|
| APOC1 | ENSG00000130208 | 19 | 45,417,504 | 45,422,606 | 13 | 7.878 | $1.67 \times 10^{-15}$ | $3.16 \times 10^{-11}$ |
| SNCA | ENSG00000145335 | 4 | 90,645,250 | 90,759,466 | 418 | 6.121 | $4.65 \times 10^{-10}$ | $8.81 \times 10^{-6}$ |
| APOE | ENSG00000130203 | 19 | 45,409,011 | 45,412,650 | 6 | 6.109 | $5.00 \times 10^{-10}$ | $9.49 \times 10^{-6}$ |
| TOMM40 | ENSG00000130204 | 19 | 45,393,826 | 45,406,946 | 53 | 6.109 | $5.00 \times 10^{-10}$ | $9.49 \times 10^{-6}$ |
| SLC50A1 | ENSG00000169241 | 1 | 155,107,820 | 155,111,329 | 5 | 5.658 | $7.66 \times 10^{-9}$ | $1.45 \times 10^{-4}$ |
| PVRL2 | ENSG00000130202 | 19 | 45,349,432 | 45,392,485 | 177 | 5.526 | $1.64 \times 10^{-8}$ | $3.11 \times 10^{-4}$ |
| MMRN1 | ENSG00000138722 | 4 | 90,800,683 | 90,875,780 | 212 | 5.096 | $1.74 \times 10^{-7}$ | $3.29 \times 10^{-3}$ |
| EFNA3 | ENSG00000143590 | 1 | 155,036,224 | 155,060,014 | 33 | 4.677 | $1.46 \times 10^{-6}$ | 0.0276 |
| EFNA3 | ENSG00000251246 | 1 | 155,036,224 | 155,059,283 | 31 | 4.668 | $1.52 \times 10^{-6}$ | 0.0289 |
| ARHGAP27 | ENSG00000159314 | 17 | 43,471,275 | 43,511,787 | 109 | 4.18 | $1.43 \times 10^{-5}$ | 0.271 |
| C1orf27 | ENSG00000157181 | 1 | 186,344,890 | 186,390,510 | 90 | 4.10 | $2.06 \times 10^{-5}$ | 0.390 |
| TPR | ENSG00000047410 | 1 | 186,280,784 | 186,344,825 | 134 | 4.09 | $2.20 \times 10^{-5}$ | 0.416 |
| PLEKHM1 | ENSG00000225190 | 17 | 43,513,266 | 43,568,115 | 151 | 3.88 | $5.33 \times 10^{-5}$ | 1.00 |
| JAKMIP3 | ENSG00000188385 | 10 | 133,918,175 | 133,998,313 | 403 | 3.84 | $6.05 \times 10^{-5}$ | 1.00 |
| STH | ENSG00000256762 | 17 | 44,076,616 | 44,077,060 | 1 | 3.78 | $7.85 \times 10^{-5}$ | 1.00 |
| MAPT | ENSG00000186868 | 17 | 43,971,748 | 44,105,700 | 725 | 3.76 | $8.54 \times 10^{-5}$ | 1.00 |
| TMEM175 | ENSG00000127419 | 4 | 926,175 | 952,444 | 69 | 3.75 | $8.84 \times 10^{-5}$ | 1.00 |
| TINAGL1 | ENSG00000142910 | 1 | 32,042,116 | 32,053,288 | 15 | 3.74 | $9.05 \times 10^{-5}$ | 1.00 |

#### Supplementary Table 7. Effect of covariate adjustment on the APOE ε4 association with neocortical versus subcortical Lewy body distribution.

| Model | N | OR | 95% CI | P |
|---|---|---|---|---|
| Primary analysis (age at death, sex, AD, PC1–PC3) | 2,309 | 1.70 | 1.45–1.98 | $2.47 \times 10^{-11}$ |
| Sex, PC1–PC3 | 1,034 | 2.30 | 1.77–2.99 | $3.9 \times 10^{-10}$ |
| with age at death | 1,034 | 2.37 | 1.82–3.09 | $1.4 \times 10^{-10}$ |
| with AD neuropathologic change | 1,034 | 2.09 | 1.60–2.72 | $6.0 \times 10^{-8}$ |
| with disease duration | 1,034 | 2.27 | 1.74–2.94 | $9.6 \times 10^{-10}$ |
| All covariates | 1,034 | 2.12 | 1.62–2.78 | $4.5 \times 10^{-8}$ |

#### Supplementary Table 8. Association of the lead variants identified in the case-control analyses with neocortical versus subcortical Lewy body distribution.

| rsID | Chr | BP | A1 | EAF | Locus | OR (95% CI) | P | I² (%) | Het P |
|---|---|---|---|---|---|---|---|---|---|
| rs2230288 | 1 | 155,236,376 | T | 0.029 | GBA1 | 1.33 (0.90–1.96) | 0.15 | 81.1[†] | 0.021 |
| rs34311866 | 4 | 958,159 | C | 0.216 | TMEM175 | 1.05 (0.91–1.22) | 0.49 | 0 | 0.836 |
| rs356182[*] | 4 | 89,704,960 | G | 0.369 | SNCA (3') | 0.97 (0.85–1.10) | 0.63 | 11.9 | 0.287 |
| rs2737030 | 4 | 89,788,948 | T | 0.832 | SNCA (5') | 1.11 (0.94–1.31) | 0.21 | 0 | 0.563 |
| rs3117443[*] | 6 | 29,384,854 | A | 0.578 | MHC | 0.90 (0.80–1.03) | 0.12 | 0 | 0.881 |
| rs9275382 | 6 | 32,701,054 | T | 0.090 | HLA class II | 1.00 (0.80–1.25) | 0.99 | 0 | 0.440 |
| rs429358 | 19 | 44,908,684 | C | 0.258 | APOE | 1.70 (1.45–1.98) | $2.47 \times 10^{-11}$ | 73.5 | 0.052 |

[†]Lead variants from the case-control analysis were extracted from the meta-analysis results before heterogeneity filtering; the I² > 80% filter was applied only to the genome-wide scan.

*From the PD/PDD-only analysis.

### Supplementary Table 9. Key Resources Table.

| RESOURCE TYPE | RESOURCE NAME | SOURCE | IDENTIFIER | NEW/REUSE |
|---|---|---|---|---|
| Dataset | Defining and Diagnosing neurodegenerative Movement Disorders through integrated analysis of Genetics and neuroPathology (MD-GAP)/Global Parkinson's Genetics Program (GP2) | AMP-PD | DOI: 10.1002/mds.28494 | NEW |
| Dataset | NIH Neurobiobank | NIHNBB | https://neurobiobank.nih.gov/ | REUSE |
| Software/code | R code: Analysis Script | Github | https://github.com/huw-morris-lab/MD-GAP-GP2-CPC.git | NEW |
| Software/code | GenoTools | https://github.com/dvitale199/GenoTools | https://doi.org/10.1093/g3journal/jkae268 | REUSE |
| Software/code | R Project for Statistical Computing, v4.3.1 | https://www.r-project.org/ | RRID:SCR_001905 | REUSE |
| Software/code | R package: dplyr | https://cran.r-project.org/web/packages/dplyr/index.html | RRID:SCR_016708 | REUSE |
| Software/code | R package: data.table | https://github.com/Rdatatable/data.table | RRID:SCR_026117 | REUSE |
| Software/code | R package: readxl | https://cran.r-project.org/web/packages/readxl/index.html | RRID:SCR_018083 | REUSE |
| Software/code | R package: stringr | https://stringr.tidyverse.org/ | RRID:SCR_022813 | REUSE |
| Software/code | R package: ggplot2 | https://cran.r-project.org/web/packages/ggplot2/index.html | RRID:SCR_014601 | REUSE |
| Software/code | R package: tidyverse | https://CRAN.R-project.org/package=tidyverse | RRID:SCR_019186 | REUSE |
| Software/code | PLINK v.2.0 | https://www.cog-genomics.org/plink/2.0/ | RRID:SCR_001757 | REUSE |
| Software/code | Julia Programming Language | https://julialang.org/ | RRID:SCR_021666 | REUSE |
| Software/code | METAL | http://csg.sph.umich.edu/abecasis/Metal/ | RRID:SCR_002013 | REUSE |
| Software/code | GCTA v.1.93.0 | https://yanglab.westlake.edu.cn/software/gcta/ | | REUSE |
| Software/code | LDSC v.1.0.1 | https://github.com/bulik/ldsc | RRID:SCR_022801 | REUSE |
| Software/code | Python Programming Language v3.10, 3.11 | http://www.python.org/ | RRID:SCR_008394 | REUSE |
| Software/code | Python: Panda | http://www.python.org/ | RRID:SCR_018214 | REUSE |
| Software/code | Python: NumPy | http://www.python.org/ | RRID:SCR_008633 | REUSE |
| Software/code | Python: SciPy | http://www.python.org/ | RRID:SCR_008058 | REUSE |
| Software/code | Python: Matplotlib | http://www.python.org/ | RRID:SCR_008624 | REUSE |

**A)**

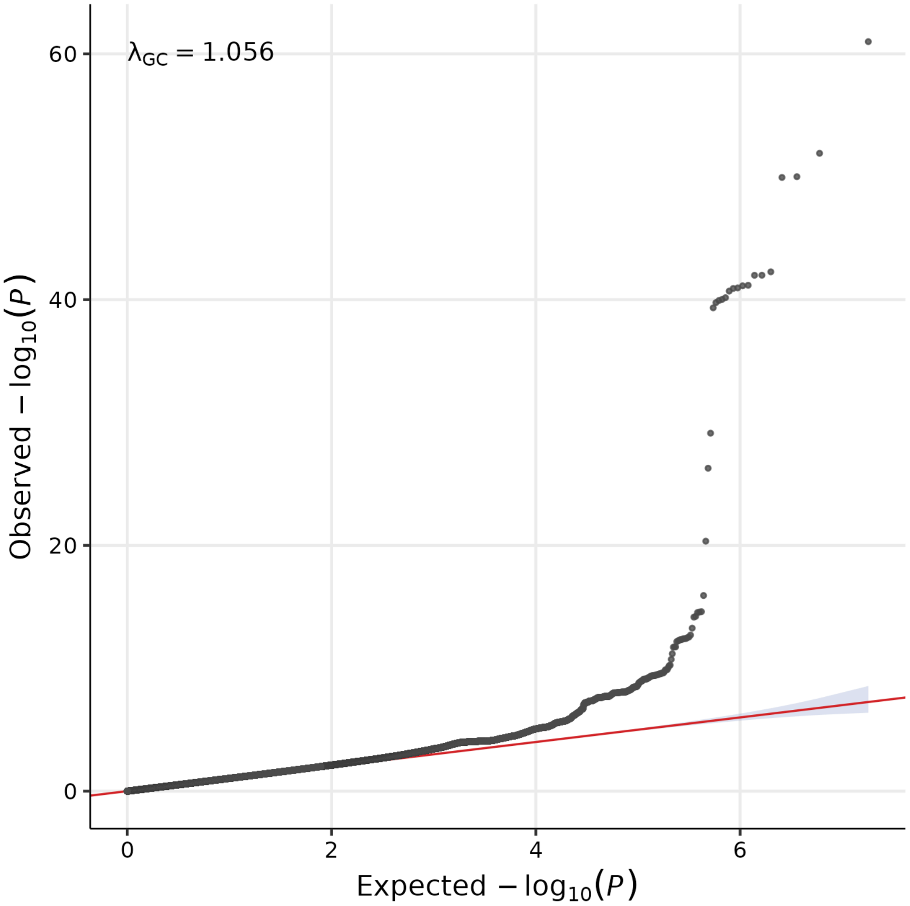

$\lambda_{GC} = 1.056$
Observed $-\log_{10}(P)$
60
40
20
0
0
2
4
6
Expected $-\log_{10}(P)$


**B)**

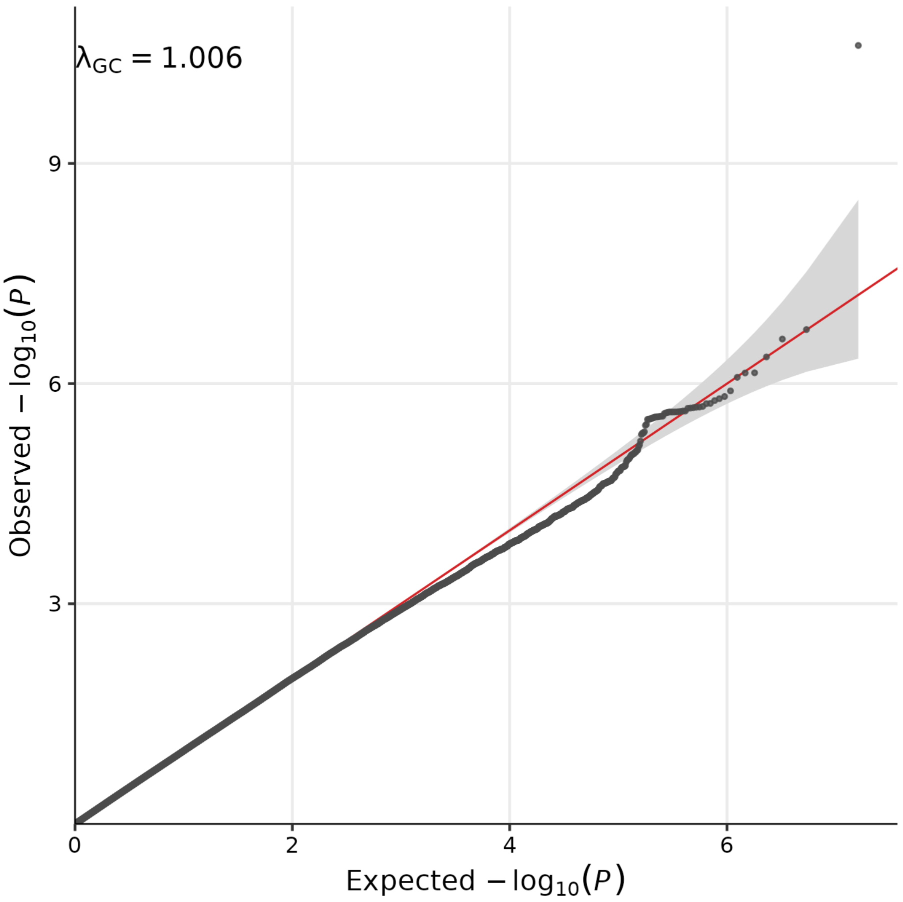


**Supplementary Figure 1. Quantile–quantile plot for the meta-analysis of autopsy-confirmed Lewy body disease.** A) Observed against expected $-\log_{10}(P)$ for 8,990,674 autosomal variants tested in 2,661 cases and 12,068 controls. The genomic inflation factor was $\lambda_{GC}$ = 1.056. B) Observed against expected $-\log_{10}(P)$ for 8,073,753 autosomal variants tested in 1,362 neocortical versus 947 subcortical cases. The genomic inflation factor was $\lambda_{GC}$ = 1.006. In both panels, the red line shows the null expectation and the shaded band its 95% confidence interval.

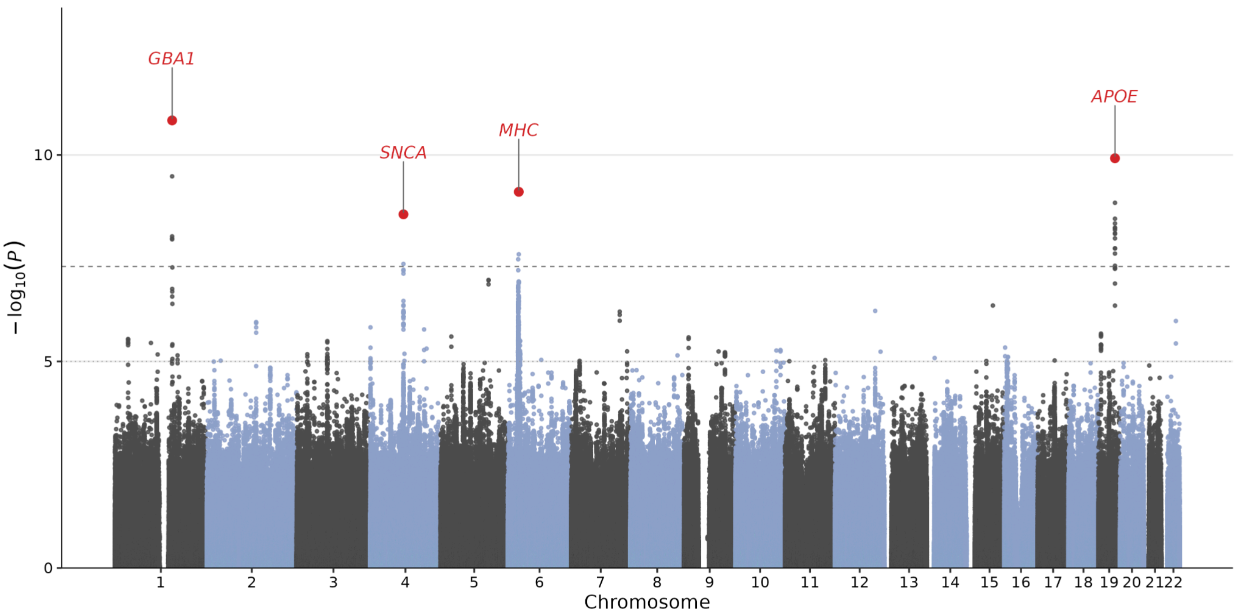


**Supplementary Figure 2. Manhattan plot of the pathologically confirmed PD/PDD-only case–control genome-wide association analysis.** Results of the PD/PDD only versus control GWAS (n = 1,216 PD/PDD cases and n = 10,517 controls). All cases had a clinical diagnosis of PD or PDD with neuropathological confirmation of LBD. Genome-wide significant loci are highlighted in red and labelled by their nearest or established candidate gene: *GBA1* (1q22), *SNCA* (4q22), *MHC* (6p21.32) and *APOE* (19q13.32). Negative logarithm p-value is represented on the y-axis, while chromosome position is represented on the x-axis. The dotted line indicates genome-wide significant threshold ($5 \times 10^{-8}$).

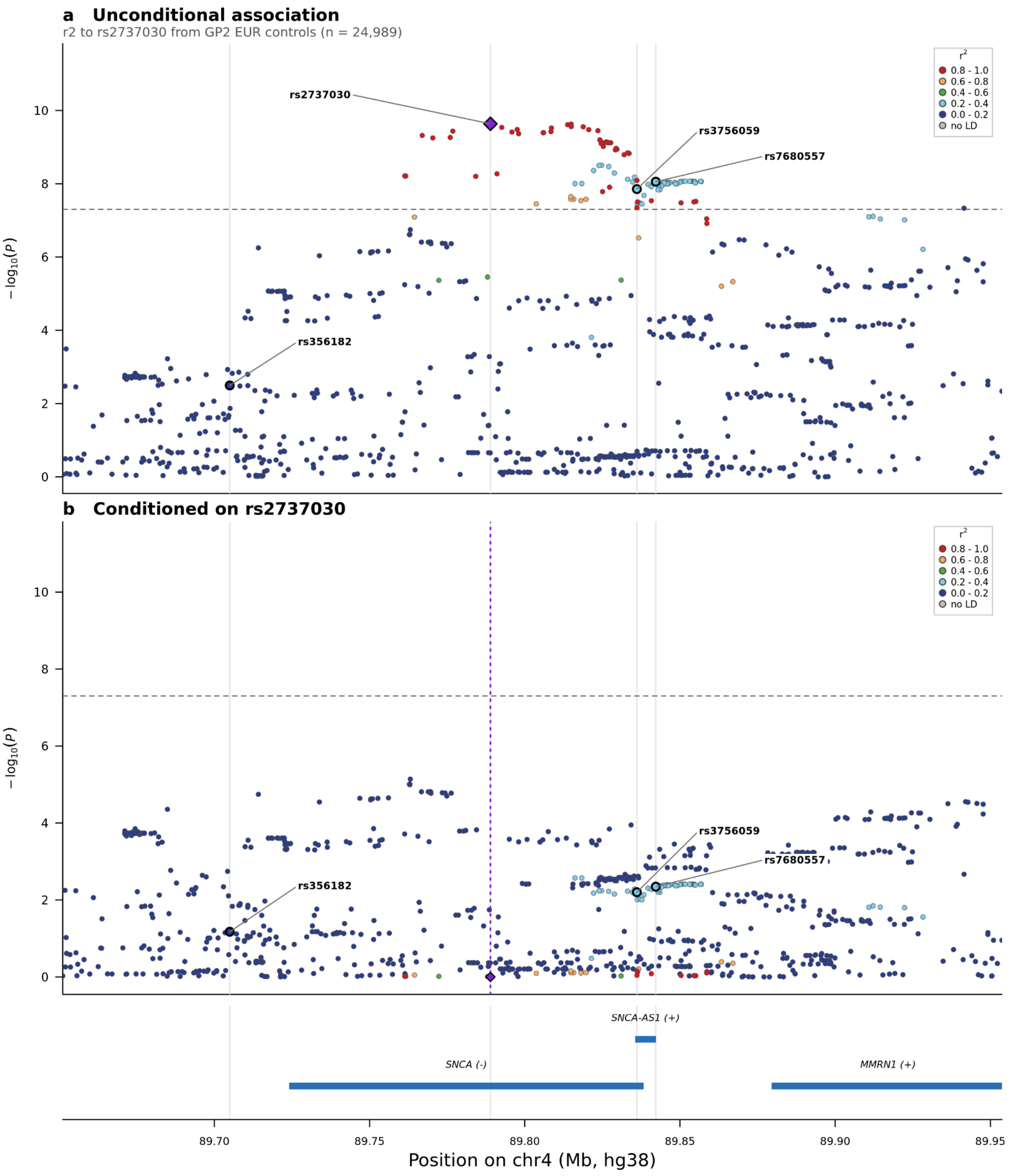


**Supplementary Figure 3. Regional association plot at the *SNCA* locus, before and after conditioning on rs2737030.** A) Regional association at the *SNCA* locus (chr4:89.65–89.95 Mb, GRCh38) plotted as a function of genomic position, denoting the index variant rs2737030 by a purple diamond. Single nucleotide variants and indels surrounding the index variant are colour-coded to reflect the strength of linkage disequilibrium with the index variant based on pairwise $r^2$-values in unrelated European-ancestry controls from GP2 (red, $1.0 \geq r^2 \geq 0.8$; orange, $0.8 > r^2 \geq 0.6$; green, $0.6 > r^2 \geq 0.4$; light blue, $0.4 > r^2 \geq 0.2$; dark blue, $0.2 > r^2 \geq 0$; grey, no $r^2$ value available). The Parkinson's disease sentinel rs356182 and the previously reported 5′ variants rs3756059 and rs7680557 are outlined in black. B) The same region conditioned on rs2737030, which is itself excluded as the conditioning variant and marked by a dotted vertical line; colour-coding and outlined variants are as in A), and the y-axis scale is shared with A). Dashed line, $P = 5 \times 10^{-8}$.

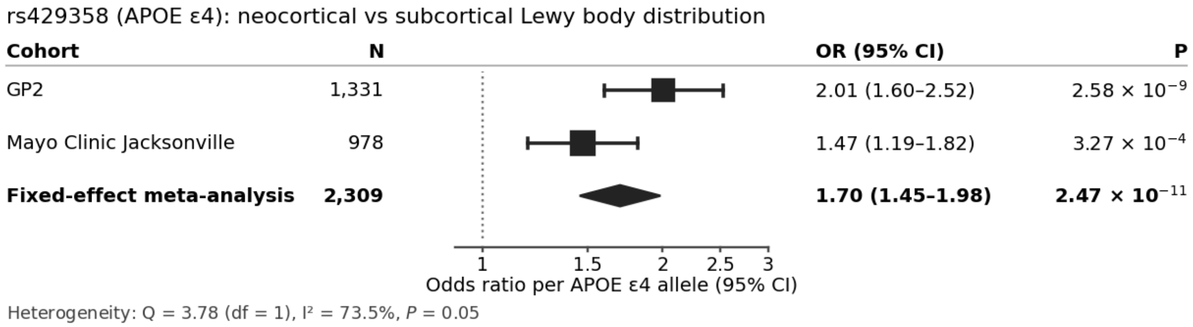


**Supplementary Figure 4. Forest plot.** Association of *APOE* ε4 (rs429358) with neocortical relative to subcortical Lewy body distribution, by cohort, after adjustment for Alzheimer's disease neuropathological change. Per-allele odds ratios are shown for the ε4-defining C allele in 2,309 autopsy-confirmed Lewy body disease cases with harmonised Lewy body staging (GP2 *n* = 1,331; Mayo Clinic Jacksonville *n* = 978), comparing cases with neocortical (*n* = 1,362) and subcortical (*n* = 947) Lewy body distribution. Models were adjusted for sex, age, Alzheimer's disease pathology and principal components 1–3.
